# The impact of fatty acids biosynthesis on the risk of cardiovascular diseases in Europeans and East Asians: *A Mendelian randomization study*

**DOI:** 10.1101/2022.04.17.22269308

**Authors:** Maria Carolina Borges, Phillip Haycock, Jie Zheng, Gibran Hemani, Laurence J Howe, A Floriaan Schmidt, James R Staley, R Thomas Lumbers, Albert Henry, Rozenn N Lemaitre, Tom R Gaunt, Michael V Holmes, George Davey Smith, Aroon D Hingorani, Deborah A Lawlor

**Author notes:** These authors contributed equally to this work (Aroon D Hingorani, Deborah A Lawlor).

## Abstract

Despite early interest, the evidence linking fatty acids to cardiovascular diseases remains controversial. We used Mendelian randomization to explore the involvement of polyunsaturated (PUFA) and monounsaturated (MUFA) fatty acids biosynthesis in the aetiology of several cardiovascular disease endpoints in up to 1,153,768 European and 212,453 East Asian ancestry individuals. As instruments, we selected single nucleotide polymorphisms (SNP) mapping to genes with well-known roles in PUFA (i.e. *FADS1/2* and *ELOVL2*) and MUFA (i.e. *SCD*) biosynthesis. Our findings suggest that higher PUFA biosynthesis rate (proxied by rs174576 near *FADS1/2*) is related to higher odds of multiple cardiovascular diseases, particularly ischemic stroke, peripheral artery disease and venous thromboembolism, whereas higher MUFA biosynthesis rate (proxied by rs603424 near *SCD*) is related to lower odds of coronary artery disease among Europeans. Results were unclear for East Asians as most effect estimates were imprecise. By triangulating multiple approaches (i.e. uni-/multi-variable Mendelian randomization, a phenome-wide scan, genetic colocalization and within-sibling analyses), our results are compatible with higher low- density lipoprotein (LDL)-cholesterol (and possibly glucose) being a downstream effect of higher PUFA biosynthesis rate. Our findings indicate that genetically-determined PUFA and MUFA biosynthesis are involved in the aetiology of cardiovascular diseases and suggest LDL-cholesterol as a potential mediating trait between PUFA biosynthesis and cardiovascular diseases risk.

## INTRODUCTION

Fatty acids constitute the main components of dietary fats and are required in human nutrition as a source of energy and for metabolic and structural activities (1). They are capable of influencing a wide range of cell signalling pathways and have been implicated in the regulation of several processes involved in the aetiology of cardiovascular diseases, including lipid metabolism (2–4), glucose homeostasis (5, 6), blood pressure (7–9), inflammatory response (10–12), and endothelial function (9, 13). Fatty acids are commonly subdivided into broad classes according to the degree of unsaturation (i.e., number of carbon- carbon double bonds) into saturated (SFA), monounsaturated (MUFA) and polyunsaturated (PUFA) fatty acids, the latter being classified as omega-3 or omega-6 PUFA depending on the position of the first double bond from the terminal methyl group.

Some fatty acids can be synthesized endogenously by fatty acid synthase or taken up by diet and further elongated and desaturated into longer chain fatty acids by fatty acid elongases and desaturases, respectively (14). Genome-wide association studies (GWAS) have reported that circulating fatty acids are strongly influenced by genetic variants near genes coding fatty acid elongases and desaturases: fatty acid desaturase 1 (*FADS1 -* ENSG00000149485), fatty acid desaturase 2 (*FADS2 -* ENSG00000134824), elongase 2 (*ELOVL2* - ENSG00000197977), and stearoyl-CoA desaturase (*SCD* - ENSG00000099194) (15–21). The chemical reactions and pathways catalysed by the enzymes encoded by *FADS1*, *FADS2*, *ELOVL2*, and *SCD* are summarised in **Figure 1**.

**Figure 1.**
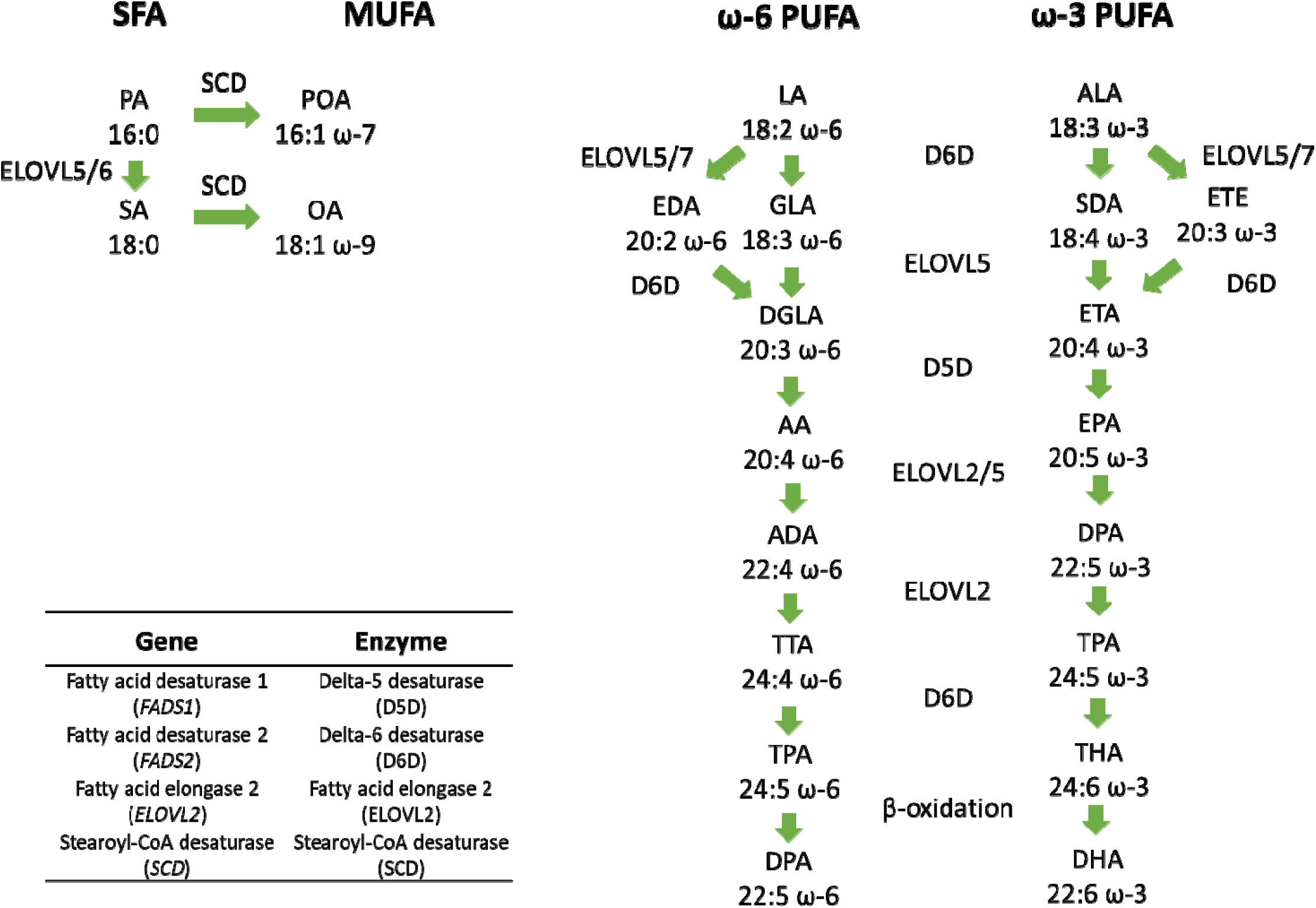
Overview of desaturation and elongation reactions involved in the conversion of MUFA from SFA and of longer-chain omega-3 and omega-6 PUFA from their shorter chain precursors. Red boxes highlight enzymes encoded by genes mapped to genetic variants strongly associated with circulating fatty acids in genome-wide association studies: delta-5 desaturase (D5D), delta-6 desaturase (D6D), fatty acid elongase 2 (ELOVL2), and stearoyl-CoA desaturase (SCD) encoded by *FADS1* (ENSG00000149485), *FADS2* (ENSG00000134824)*, ELOVL2* (ENSG00000197977), *SCD* (ENSG00000099194), respectively. *Saturated fatty acids (SFA):* PA: palmitic acid; SA: stearic acid. *Monounsaturated fatty acids (MUFA):* POA: palmitoleic acid; OA: oleic acid. ω*-6 polyunsaturated fatty acids (PUFA):* LA: linoleic acid; GLA: γ-linolenic acid; EDA: eicosadienoic acid; DGLA: dihomo-γ-linolenic acid; AA: arachidonic acid; ADA: adrenic acid; TTA: tetracosatetraenoic acid; TPA: tetracosapentaenoic acid; DPA: docosapentaenoic acid. ω*-3 polyunsaturated fatty acids (PUFA):* ALA: α-linolenic acid; SDA: stearidonic acid; ETE: eicosatrienoic acid; ETA: eicosatetraenoic acid; EPA: eicosapentaenoic acid; DPA: docosapentaenoic acid; TPA: tetracosapentaenoic acid; THA: tetracosahexaenoic acid; DHA: docosahexaenoic acid.

Mendelian randomization uses genetic variants associated with putative risk factors as instruments to assess their involvement in disease aetiology (22–24). The use of human genetics to explore the effect of modifiable risk factors on cardiometabolic diseases, such as in Mendelian randomization, has proven valuable to (de)prioritise targets for intervention and to assess potential target-mediated adverse effects reducing late-stage failures in RCTs due to lack of efficacy or from target-mediated adverse reactions (25).

Genetic variants affecting the expression or activity of genes encoding for fatty acid elongases and desaturases (e.g. *FADS1/2*, *ELOVL2*, and *SCD*) can be used as causal anchors in Mendelian randomization studies investigating the involvement of fatty acids in the development of cardiovascular diseases. Most previous Mendelian randomization studies investigating the role of fatty acids on the risk of cardiovascular diseases have relied solely or heavily on genetic variants within the *FADS1/2* locus involved in PUFA synthesis by encoding the enzymes delta-5 desaturase (D5D) and delta-6 desaturase (D6D), respectively. Overall, these studies have reported that shorter (e.g. α-linolenic acid (ALA) and linoleic acid (LA)) and longer (e.g. arachidonic acid (AA)) chain PUFA are associated with lower and higher risk of cardiovascular diseases, respectively (26–32).

These studies potentially strengthen the evidence on the involvement of fatty acids in the development of cardiovascular diseases given the well-established link of D5D/D6D with PUFA biosynthesis. However, such studies suffer from a critical limitation given *FADS1/2* variants are not reliable instruments for individual fatty acids. First, *FADS1/2* variants will affect multiple fatty acids on the same pathway and, in some cases, on different pathways with reactions catalysed by D5D/D6D (**Figure 1**). Second, these studies have not extensively explored whether the association of *FADS1/2* variants with cardiovascular diseases risk could be explained by biological pathways independent of fatty acids (e.g. if variants simultaneously influence the expression of other genes in the region that affect cardiovascular diseases) or due to confounding by population stratification or other familial mechanisms (e.g., indirect genetic effects).

The aim of this study was to use Mendelian randomization to explore the effect of fatty acids biosynthesis on a wide range of cardiovascular disease end-points in up to 1,153,768 European and 212,453 East Asian ancestry individuals. We extend work in previous studies by using genetic variants regulating multiple rate-limiting enzymes in fatty acids biosynthesis (i.e. D5D/D6D, ELOVL2 and SCD), comparing findings between Europeans and East Asians and extensively exploring the key scenarios that could lead to spurious findings in this and previous Mendelian randomization studies.

## MATERIAL AND METHODS

### Study design

We used two-sample Mendelian randomization to probe the lifelong effect of fatty acids biosynthesis on the risk of multiple cardiovascular diseases in Europeans and East Asian individuals. As instruments, we selected genetic variants mapping to genes with a well-known role in fatty acids biosynthesis (i.e. *FADS1/2*, *ELOVL2*, and *SCD*). We used two approaches to circumvent limitations in previous studies.

First, we used genetic variants to instrument for enzyme activity in a given fatty acids biosynthesis pathway (rather than for individual fatty acids) by deriving the ratio between fatty acids that are the product and the substrate of a reaction catalysed by the corresponding enzyme. This allows harnessing the advantages of cis-acting variants in the vicinity of genes coding for key enzymes in fatty acids biosynthesis pathways and can provide more credible evidence on the likely therapeutic benefit of targeting such protein in preventing cardiovascular diseases(33).

Second, we used a range of methods to assess the plausibility of the four key scenarios that could invalidate inferences from this and previous studies: (i) horizontal pleiotropy, where the genetic variant influences the outcome via a different biological pathway; (ii) confounding by *linkage disequilibrium* (LD), where the selected genetic variant is in LD with another genetic variant influencing the outcome independently; (iii) confounding by population stratification, assortative mating or indirect genetic effects, which could create a spurious association between genetic variant and outcome, and (iv) selection bias, where the genetic variant (or, more likely, its downstream traits) and the outcome affect selection into the sample resulting in a spurious association (**Figure 2**).

**Figure 2.**
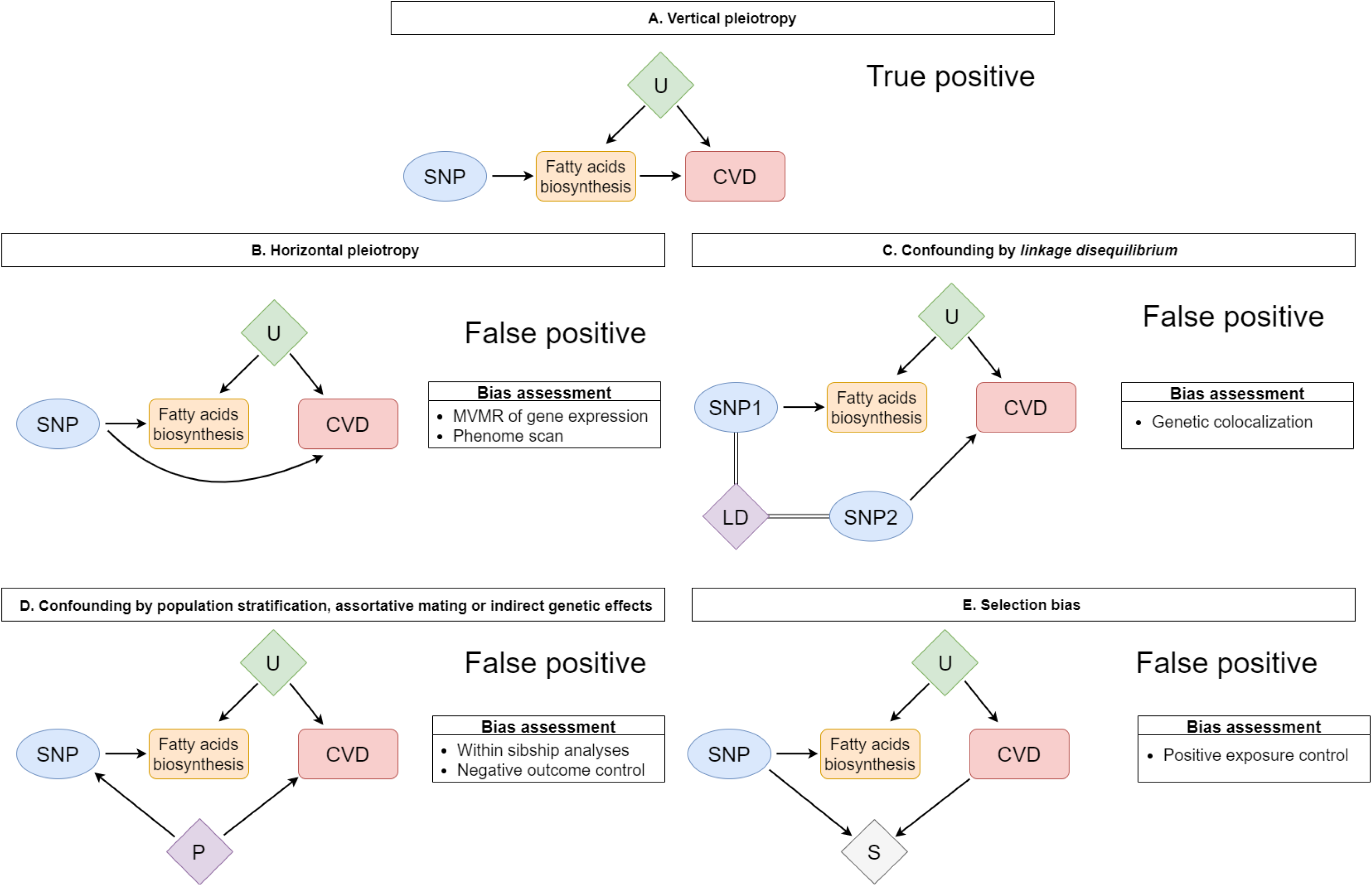
Schematic representation of multiple putative mechanisms underlying the association of genetic variants with fatty acids biosynthesis and cardiovascular disease (CVD) risk Figure 2A represents vertical pleiotropy, in which the effect of genetic instruments on CVD is mediated by fatty acids biosynthesis. Figures 2B to 2E represent alternative mechanisms that could bias Mendelian randomization findings: horizontal pleiotropy (2B), in which the genetic variant influences fatty acids biosynthesis and CVD via two different biological pathways, (2C) confounding by *linkage disequilibrium* (LD), in which the selected genetic variant is in LD with another genetic variant influencing CVD independently, (2D) confounding, in which different phenomena can introduce spurious association between genetic variant and CVD in samples of unrelated individuals, and (2D) selection bias, in which selection into the study creates a spurious association between genetic variant and CVD due to collider stratification bias. MVMR: multivariable Mendelian randomization.

### Data sources

The study included data from multiple consortia of genetic association studies (34–38) and biobanks (39–45).

#### Genetic associations with cardiovascular disease outcomes

The outcomes of interest were (prevalent/incident) coronary artery disease, ischemic stroke, haemorrhagic stroke, heart failure, atrial fibrillation, peripheral arterial disease, aortic aneurysm, venous thromboembolism, and aortic valve stenosis.

Summary data for the association between genetic variants and these cardiovascular disease endpoints was obtained from UK Biobank, FinnGen, BioBank Japan and several large-scale genetic consortia of cardiovascular disease outcomes. If genetic association data on a cardiovascular endpoint were available from two or more independent datasets of individuals from the same genetic ancestry (i.e. UK Biobank and FinnGen), genetic association estimates were pooled across data sources using fixed-effect meta-analysis with inverse variance weights. Characteristics of studies and criteria for case definition are detailed in **Supplementary table 1** and **Supplementary methods**.

For individuals of European ancestry only/predominantly (i.e. UK Biobank, FinnGen and large-scale genetic consortia), data were available on all outcomes of interest: coronary artery disease (N cases/controls = 123,668/702,156), ischemic stroke (N cases/controls = 53,395/1,030,253), haemorrhagic stroke (N cases/controls = 4,558/627,188), heart failure (N cases/controls = 64,696/1,089,072), atrial fibrillation (N cases/controls = 77,945/1,067,430), peripheral arterial disease (N cases/controls = 9,836/627,950), aortic aneurysm (N cases/controls = 9,735/730,073), venous thromboembolism (N cases/controls = 25,284/616,235), and aortic valve stenosis (N cases/controls = 2,844/461,776).

For individuals of East Asian ancestry (i.e. BioBank Japan), cardiovascular outcomes data were available for coronary artery disease (N cases/controls = 29,319/183,134), ischemic stroke (N cases/controls = 17,671/192,383), haemorrhagic stroke (N cases/controls = 2,820/192,383), heart failure (N cases/controls = 9,413/203,040), atrial fibrillation (N cases/controls = 8,180/28,612), and peripheral arterial disease (N cases/controls = 3,593/208,860).

#### Genetic associations with circulating fatty acid concentration

For European ancestry individuals, we used genetic association data on circulating fatty acids from *The Cohorts for Heart and Aging Research in Genomic Epidemiology* (CHARGE) consortium, which has high resolution profiling of circulating fatty acids (N = 26 fatty acids measures) measured in 8,631-8,866 individuals (15–17). We also used data from two other genetic association meta-analyses on circulating fatty acids (18, 19) for assessing replication as detailed in ‘Data analysis’ under ‘Assessing the impact of genetic instruments on the fatty acids pool’.

For East Asian ancestry individuals, we used genetic association data on fatty acids from the Singapore Chinese Health Study (SCHS) for circulating PUFA (N = 1,361) (21) and from a metanalysis of the Nutrition and Health of Aging Population in China (NHAPC) and the Chinese ancestry individuals of the Multi-Ethnic Study of Atherosclerosis (MESA) for RBC or circulating SFA and MUFA (N = 3,521) (46, 47).

Characteristics of these studies are detailed in **Supplementary table 2.**

### Data analysis

#### Selection of genetic instruments indexing fatty acids biosynthesis

We selected genetic variants mapping to genes that have well-characterised roles in fatty acids biosynthesis and have been previously reported by GWAS to influence circulating fatty acids (**Figure 1**). In Europeans, three genomic regions were eligible, harbouring *FADS1/2*, *ELOVL2* and *SCD* genes, whereas, in East Asians, only the *FADS1/2* locus was strongly associated with circulating fatty acids, which may be related to the modest sample size available for East Asians (N = 1,361-3,521). *FADS1/2* were considered as one single genomic region since these genes are in close proximity to each other (i.e. 0.8 kb) on the long arm of human chromosome 11.

Genetic variants regulating the expression/activity of *FADS1/2*, *ELOVL2* and *SCD* will affect multiple fatty acids on the same pathway and, in some cases, on different pathways with reactions catalysed by the same enzymes (**Figure 1**). As a result, selecting genetic variants for individual fatty acids can be highly redundant. Instead, we selected the genetic variant (± 500 kB of the target gene) most strongly related (p-value < 5 x:f 10^-8^) to a proxy of the enzyme activity (i.e. the ratio between fatty acids that are the product and the substrate of a reaction catalysed by a particular enzyme) within each genomic locus (details in **Supplementary methods**). As an example, a higher ratio of AA to dihomo-γ-linoleic acid (DGLA) would indicate more active conversion due to higher expression/activity of D5D, the enzyme encoded by *FADS1*.

For European ancestry individuals, we derived genetic association data for proxies of enzyme activity by applying the GWIS (“Genome-wide Inferred Study”) method to genetic association data for circulating fatty acids from the CHARGE consortium (15–17) for the ratios of AA to DGLA (proxy of D5D activity), γ-linolenic acid (GLA) to LA (proxy of D6D activity), docosahexaenoic acid (DHA) to docosapentaenoic acid (DPA n-3) (proxy of ELOVL2 activity), and palmitoleic acid (POA) to palmitic acid (PA) (proxy of SCD activity), as detailed in **Supplementary methods**. Briefly, GWIS approximates GWAS summary statistics for a new variable as a function of multiple phenotypes for which GWAS summary statistics, phenotypic means, and covariances are available (48).

For East Asian ancestry individuals, the original GWAS investigators derived genetic association data for the proxies of enzyme activity from individual level data on circulating fatty acids (21), as follows: AA to DGLA (proxy of D5D activity) and DGLA to LA (proxy of D6D activity).

If the selected genetic variant was a palindromic SNP, it was replaced by a non-palindromic proxy variant in strong LD to avoid data harmonisation problems in subsequent analyses. All SNP-trait associations were harmonised so that the allele associated with increasing enzyme activity was the effect allele, indicating more active conversion. We approximated the R^2^, a measure of the variance in exposure explained by the genetic variant, and the F-statistics, a measure of instrument strength (49), as detailed in **Supplementary methods**.

#### Assessing the impact of genetic instruments on the fatty acids pool

We assessed the impact of the selected genetic instruments on the circulating fatty acids pool in the discovery samples (i.e. CHARGE in Europeans and SCHS in East Asians) for internal validation. Among European ancestry individuals, we could test for replication in two independent datasets (external validation) (18, 19).

#### Mendelian randomization analysis

For each cardiovascular outcome, we used the Wald ratio method to estimate the odds ratio of disease per standard unit increase in enzyme activity by dividing estimates for the genetic association with cardiovascular outcome by estimates for the genetic association with enzyme activity as detailed in the **Supplementary methods**. We used a Bonferroni correction to account for the number of outcomes available in Europeans (p-value = 0.05/9 outcomes = 0.006) and East Asians (p-value = 0.05/6 outcomes = 0.008). We use these p-value thresholds simply as a heuristic for highlighting associations worthy of follow-up. Mendelian randomization analyses were performed using R software version 3.6.2 (R Foundation for Statistical Computing) including the TwoSampleMR R package (50).

#### Exploring bias in Mendelian randomization analyses

We explored the validity of our Mendelian randomization findings by assessing whether results are likely to be spurious due to horizontal pleiotropy, population stratification, assortative mating, indirect genetic effects, and selection bias as follows.

##### Horizontal pleiotropy

Horizontal pleiotropy is one of the main threats to the validity of Mendelian randomization studies since it is a widespread biological phenomenon (51) and it cannot be empirically verified. We combined two approaches to explore the plausibility that our results are explained by horizontal pleiotropy: phenome scan and multivariable Mendelian randomization to adjust for the potential effect of other genes expressed in the candidate genomic regions.

###### Phenome scan

We explored the potential mechanisms that might link the selected genetic variants to cardiovascular diseases by testing their association with well-established cardiovascular risk factors (hypothesis-driven approach) and across the phenome (hypothesis-free approach) (52).

In the hypothesis-driven approach, we tested for the association between the selected genetic variants and eight well-established risk factors for cardiovascular diseases [i.e. low-density lipoprotein (LDL)-cholesterol, triglycerides, systolic, diastolic blood pressure, fasting glucose, type 2 diabetes, smoking, and body mass index] using Bonferroni correction to account for multiple testing (p-value = 0.05/8 risk factors). Genetic association data for these risk factors were extracted from UK Biobank (only/predominantly) and BioBank Japan using the IEU OpenGWAS project database (53).

In the hypothesis-free (phenome-wide) approach, we used an automated phenome-wide scan tool from the IEU OpenGWAS project database (53) to test the association of the selected genetic variants with 32,534-34,465 (non-unique) traits for European ancestry individuals and 110 traits for Japanese individuals from BioBank Japan. We used a Bonferroni correction to account for multiple testing considering the maximum number of traits included in the phenome-wide scan in Europeans (p-value = 0.05/34,465 = 1.5 x 10^-6^) and East Asians (p-value = 0.05/110 = 4.5 x 10^-4^).

###### Gene expression and tissue-specific analyses

We explored the influence of higher expression of the target genes (i.e. *FADS1/2*, *ELOVL2* and *SCD*), and their tissue specificity, on cardiovascular disease risk and risk factors by integrating expression quantitative trait loci (eQTL) data from Genotype-Tissue Expression (GTEx) version 8 with genetic association data for cardiovascular traits, as detailed in **Supplementary methods**. First, we performed a cross-tissue assessment of the association of the selected genetic variants with transcription of cis-genes, defined as genes for which the transcription start site was 1 Mb away from the genetic variant. Second, we used multivariable Mendelian randomization to jointly model the expression of a target gene (i.e. *FADS1/2*, *ELOVL2*, and *SCD*) and a co-expressed cis-gene on cardiovascular outcomes across tissues using the MVMR R package (54). This analysis allowed us to estimate the direct contribution of changes in the expression of each target gene where the selected genetic variant was related to co-expression of a non-target cis-gene.

##### Confounding by linkage disequilibrium

*Linkage disequilibrium* (LD) could lead to spurious results in Mendelian randomization analysis if, by chance, the selected genetic variant influencing fatty acids biosynthesis is correlated (i.e. in LD) with another genetic variant influencing the risk of cardiovascular diseases independently. We used *coloc* (55), a method for pairwise genetic colocalization analysis, to test whether the same genetic variant influences fatty acids biosynthesis and cardiovascular diseases risk. A posterior probability of association (PPA) ≥ 70% for association with both traits due to a single causal variant was considered as strong evidence for a shared genetic variant.

*Coloc* assumes a single causal variant in the genomic region, and, as a result, the presence of multiple conditionally independent SNPs within a region can affect the performance of the method. Therefore, where *Coloc* provided evidence of a shared genetic signal, we also performed approximate conditional analyses using GCTA(56, 57) (adjusting for the top SNP in each genomic region) and re-ran *Coloc* using the adjusted association estimates as a sensitivity analyses.

Colocalization analyses were restricted to European datasets as the method assumes that samples are drawn from independent populations of similar genetic background (i.e. allele frequencies and LD pattern are identical), which was not the case for East Asians in our analyses since fatty acids and cardiovascular disease data were derived from Singaporean Chinese and Japanese individuals, respectively.

##### Population stratification, assortative mating and indirect genetic effects

Mendelian randomization studies generally assume that genetic association estimates from GWAS reflect the direct effect of a genetic variant on a phenotype, i.e., the downstream effect of inheriting an allele. However, there is growing evidence that GWAS of unrelated individuals may also capture non-direct sources of association relating to population stratification, assortative mating and indirect genetic effects of parents(58). Within-family GWAS/Mendelian randomization designs, such as parent-offspring trio or within-sibship models, control for variation in parental genotypes, and so are not affected by these potential biases (59–61).

There is evidence that the *FADS1/2* locus was under important selection pressure in different populations and at different times, possibly as a response to dietary changes and the need for adequate supply of essential long-chain PUFA from precursors (62). Despite attempts to control for population stratification in genetic association data (e.g. by adjusting for genomic principal components), there could still be residual population structure as has recently been shown in UK Biobank (63). In addition, there is a possibility that indirect genetic effects of parents bias studies among unrelated individuals given the literature suggesting that maternal genotype for *FADS1/2* variants might indirectly influence offspring outcomes via intrauterine effects and/or breastfeeding (64).

We used two approaches to explore this. First, we tested the association between the selected genetic variants with two negative control outcomes, skin colour and ease of skin tanning, using UK Biobank genetic association data deposited in the IEU Open GWAS Project (53). Since these traits could not conceivably be affected by fatty acids biosynthesis, any evidence for an association between genetic variants and these negative control outcomes would be indicative of residual population stratification(65). Second, we used data from a recent within-sibship GWAS, including up to 178,076 individuals (77,832 sibling pairs) from 23 cohorts, to evaluate if our findings are sensitive to population stratification, assortative mating, and indirect genetic effects of parents. We compared the within-sibship association of the selected genetic variants with cardiovascular risk factors (LDL-cholesterol, triglycerides, systolic blood pressure, glycated haemoglobin, smoking, and body mass index) with estimates from standard GWAS models in unrelated individuals (sample size ranging from 50,361 for glycated haemoglobin to 155,457 for BMI). Data on cardiovascular disease endpoints and other risk factors (i.e. diastolic blood pressure, fasting glucose, and type 2 diabetes) were not available.

##### Selection bias

Several processes of sample selection, occurring from study design to data analyses, can result in selected samples not representative of their target populations (e.g. due to case/control ascertainment, participant dropout, loss to follow-up, subgroup analysis, or missing data). In some instances, non-random sample selection can bias inference about the causal effect of an exposure on an outcome, including when using Mendelian randomization (66).

Although bias can result from multiple selection mechanisms, we were particularly concerned about selection due to ascertainment of cardiovascular disease status given the design of the studies used for cardiovascular disease endpoints. As an example, BioBank Japan is a hospital-based study, in which cases for cardiovascular diseases, except atrial fibrillation, were compared to a control group including a mixture of hospital-based (i.e. individuals diagnosed at health centres with other diseases) and community-based (i.e. individuals from population-based cohorts) controls as previously described (42). In addition, UK Biobank has a response rate of 5.5% and its participants have fewer self-reported health conditions and are more likely to be older, female, wealthier, leaner, non-smokers, non-drinkers than the general UK population (67).

To explore whether these processes of sample selection could bias our findings, we adopted a positive exposure control approach in which we used Mendelian randomization to estimate the effect of well-established cardiovascular risk factors (i.e. LDL-cholesterol, triglycerides, systolic, diastolic blood pressure, glucose, type 2 diabetes, smoking, and body mass index) on the risk of cardiovascular diseases. If the effects estimated in the positive control analyses were compatible with what expected and were comparable across data sources, such analyses would argue against selection being a major source of bias.

## RESULTS

### Selection of genetic instruments indexing fatty acids biosynthesis

In individuals of European ancestry, the selected genetic variants were rs174546 (*FADS1/2*, chr11q13.3), rs174576 (*FADS1/2*, chr11q13.3), rs3734398 (*ELOVL2*, chr6q15), and rs603424 (*SCD*, chr10q22.1), which explained a proportion of the variance in the corresponding marker of enzyme activity of 32.6% (F = 4174) for AA:DGLA, 6.3% (F = 580) for GLA:LA, 2.4% (F = 218) for DHA:n-3 DPA, and 1.1% (F = 100) for POA:PA, respectively (**S**upplementary table 3). The *FADS1/2* SNPs (i.e. rs174546 and rs174576) were in strong LD (R^2^ = 0.93 1000 Genomes European population), and, therefore, only the SNP more strongly associated with the corresponding marker of enzyme activity was used in subsequent analyses (i.e. rs174546).

In individuals of East Asian ancestry, the top variant in the *FADS1/2* locus was palindromic and, therefore, was replaced by rs174546 (i.e. LD R^2^ = 0.93 in 1000G East Asian population), which explained 8.4% (F = 125) of the variance in DGLA:LA (**Supplementary table 3**). No genetic variants were associated with markers of D5D activity (AA:DGLA). As previously mentioned, genetic variants at *ELOVL2* and *SCD* loci were not associated with circulating fatty acids in East Asians and, therefore, were not eligible for our analyses, which may be related to the modest sample size available for East Asians (N = 1,361-3,521).

### Impact of genetic instruments on circulating fatty acids

Overall, the effect of genetic variants on the fatty acids pool was replicable across studies and ancestries. In addition, the genetic variants impact on the fatty acids pool was consistent with their predicted function on fatty acids biosynthesis. The *FADS1/2* SNP (rs174546) was associated with a lower concentration of shorter chain omega-3 (e.g. ALA) and omega-6 (e.g. LA) fatty acids and higher concentration of longer chain omega-3 (e.g. DHA) and omega-6 (e.g. AA) fatty acids. The *ELOVL2* SNP (rs3734398) was mostly associated with higher concentration of DHA and lower concentration of eicosapentaenoic acid (EPA) and n-3 DPA, whereas the *SCD* SNP (rs603424) was related to lower SFA, particularly PA, and higher monounsaturated fatty acids (MUFA), particularly POA (**Supplementary figures 1 and 2**).

### Mendelian randomization analysis

In European ancestry individuals, higher *FADS1*/D5D activity (proxied by increase in AA:DGLA in standard units) was related to higher odds of coronary artery disease (OR = 1.02; 95% CI: 1.01, 1.03; p-value = 0.006), ischemic stroke (OR = 1.03; 95% CI: 1.01, 1.05; p-value = 0.004), heart failure (OR = 1.02; 95% CI: 1.01, 1.04; p-value = 0.008), atrial fibrillation (OR = 1.02; 95% CI: 1.00, 1.03; p-value = 0.04), peripheral artery disease (OR = 1.08; 95% CI: 1.04, 1.12; p-value = 1 x 10^-5^), venous thromboembolism (OR = 1.07; 95% CI: 1.05, 1.09; p-value = 5 x 10^-9^), and aortic valve stenosis (OR = 1.08; 95% CI: 1.01, 1.15; p-value = 0.02). Only results for ischemic stroke, peripheral artery disease, and venous thromboembolism passed our threshold for multiple testing correction (P < 0.006) (**Figure 3**). Overall, results were consistent across studies, except for coronary artery disease, for which the estimated effect was attenuated in UK Biobank compared to other studies, and for aortic aneurysm, for which the estimated effect was in different directions between UK Biobank and other studies (**Supplementary figure 3**).

**Figure 3.**
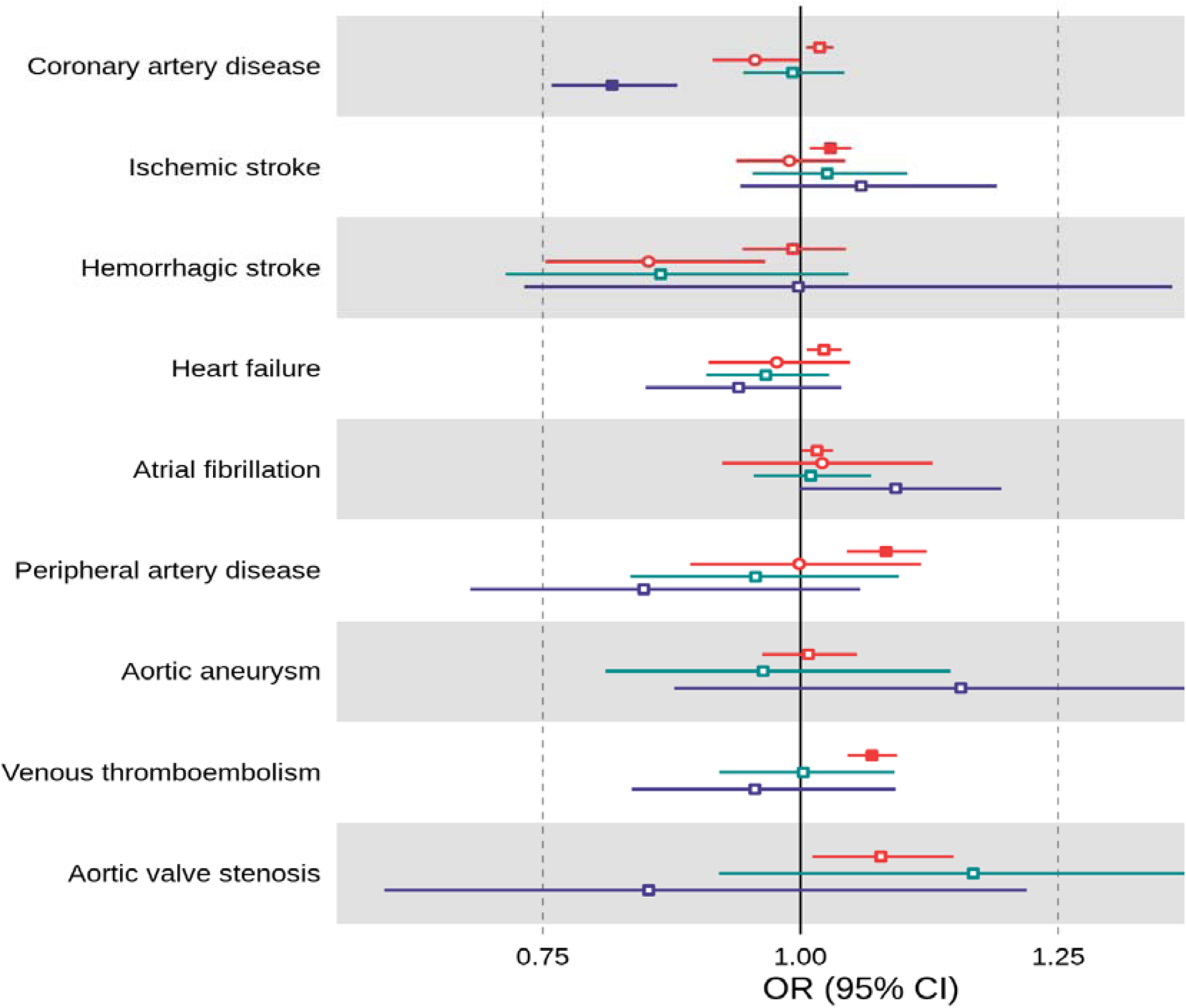
Mendelian randomization results for the risk of cardiovascular diseases related to increasing activity of enzymes coded by *FADS1/2* (D5D/D6D), *ELOVL2* (ELOVL2) and *SCD* (SCD) among individuals of European and East Asian ancestries. Results are expressed as odds ratio of cardiovascular diseases per standard unit increase in the marker of enzyme activity for *FADS1/2* (i.e. AA:DGLA ratio in Europeans and DGLA:LA ratio in East Asians), *ELOVL2* (i.e. DHA:DPA ratio in Europeans) and *SCD* (i.e. POA:PA ratio in Europeans) loci. For individuals of European ancestry, SNP-cardiovascular diseases association data were metanalysed across multiple genetic association consortia, UK Biobank and FinnGen. For individuals of East Asian ancestry, SNP-cardiovascular diseases association data were extracted from BioBank Japan. Full symbols indicate associations at P-value lower than the P-value threshold accounting for multiple testing (P < 0.006 for Europeans and P < 0.008 for East Asians). AA: arachidonic acid; DGLA: dihomo-γ-linolenic acid; DHA: docosahexaenoic acid; DPA: docosapentaenoic acid; LA: linoleic acid; PA: palmitic acid; POA: palmitoleic acid; SNP: single nucleotide polymorphism; *FADS1/2*: fatty acids desaturase 1/2; *ELOVL2*: elongase 2; *SCD*: stearoyl-CoA desaturase.

There was little evidence supporting a relationship between higher *ELOVL2*/ELOVL2 activity (proxied by increase in DHA:DPAn-3 in standard units) and cardiovascular endpoints. However, some results were imprecisely estimated and, therefore, we cannot rule out the presence of clinically meaningful effects, particularly for haemorrhagic stroke (OR = 0.86; 95% CI: 0.71, 1.05; p-value = 0.14) and aortic valve stenosis (OR = 1.17; 95% CI: 0.92, 1.48; p-value = 0.20) (**Figure 3** and **Supplementary figure 3**).

Higher *SCD*/SCD activity (proxied by increase in POA:PA in standard units) was related to lower odds of coronary artery disease (OR = 0.82; 95% CI: 0.76, 0.88; p-value = 1x 10^-7^) (**Figure 3**), which was consistent across studies (**Supplementary figure 3**). There was little evidence supporting a relationship between higher *SCD*/SCD activity (proxied by increase in POA:PA in standard units) and other cardiovascular endpoints. However, some of these results were imprecisely estimated and, therefore, we cannot rule out the presence of clinically meaningful effects, particularly for peripheral artery disease (OR = 0.85; 95% CI: 0.68, 1.06; p-value = 0.14), aortic aneurysm (OR = 1.16; 95% CI: 0.88, 1.52; p-value = 0.30), and aortic valve stenosis (OR = 0.85; 95% CI: 0.60, 1.22; p-value = 0.38) (**Figure 3** and **Supplementary figure 3**).

In East Asian ancestry individuals, there was limited evidence supporting a relationship between *FADS2*/D6D activity and the odds of cardiovascular endpoints. However, statistical power was substantially lower for analyses in East Asian individuals and, therefore, some findings could be compatible with higher *FADS2/D6D* activity (proxied by increase in DGLA:LA in standard units) being related to higher odds of disease, such as for atrial fibrillation (OR = 1.02; 95% CI: 0.92, 1.13; p-value = 0.68), or to lower odds of diseases, such as for coronary artery disease (OR = 0.96; 95% CI: 0.91, 1.00; p-value = 0.04) and haemorrhagic stroke (OR = 0.85; 95% CI: 0.75, 0.97; p-value = 0.01) (**Figure 3**).

### Exploring bias in Mendelian randomization analyses

#### Horizontal pleiotropy

##### Phenome scan

In a hypothesis-driven approach, higher D5D/D6D (*FADS1/2*) activity was related to higher LDL-cholesterol, fasting glucose, and type 2 diabetes risk, but lower triglycerides and diastolic blood pressure, among individuals of European (as proxied by AA:DGLA) and East Asian ancestry (as proxied by DGLA:LA). Higher *ELOVL2*/ELOVL2 activity (proxied by DHA:DPAn-3) was not related to cardiovascular risk factors at p-value < 0.00625 and higher *SCD*/SCD activity (proxied by POA:PA) was related to lower LDL-cholesterol, triglycerides, systolic and diastolic blood pressure (**Figure 4**);

**Figure 4.**
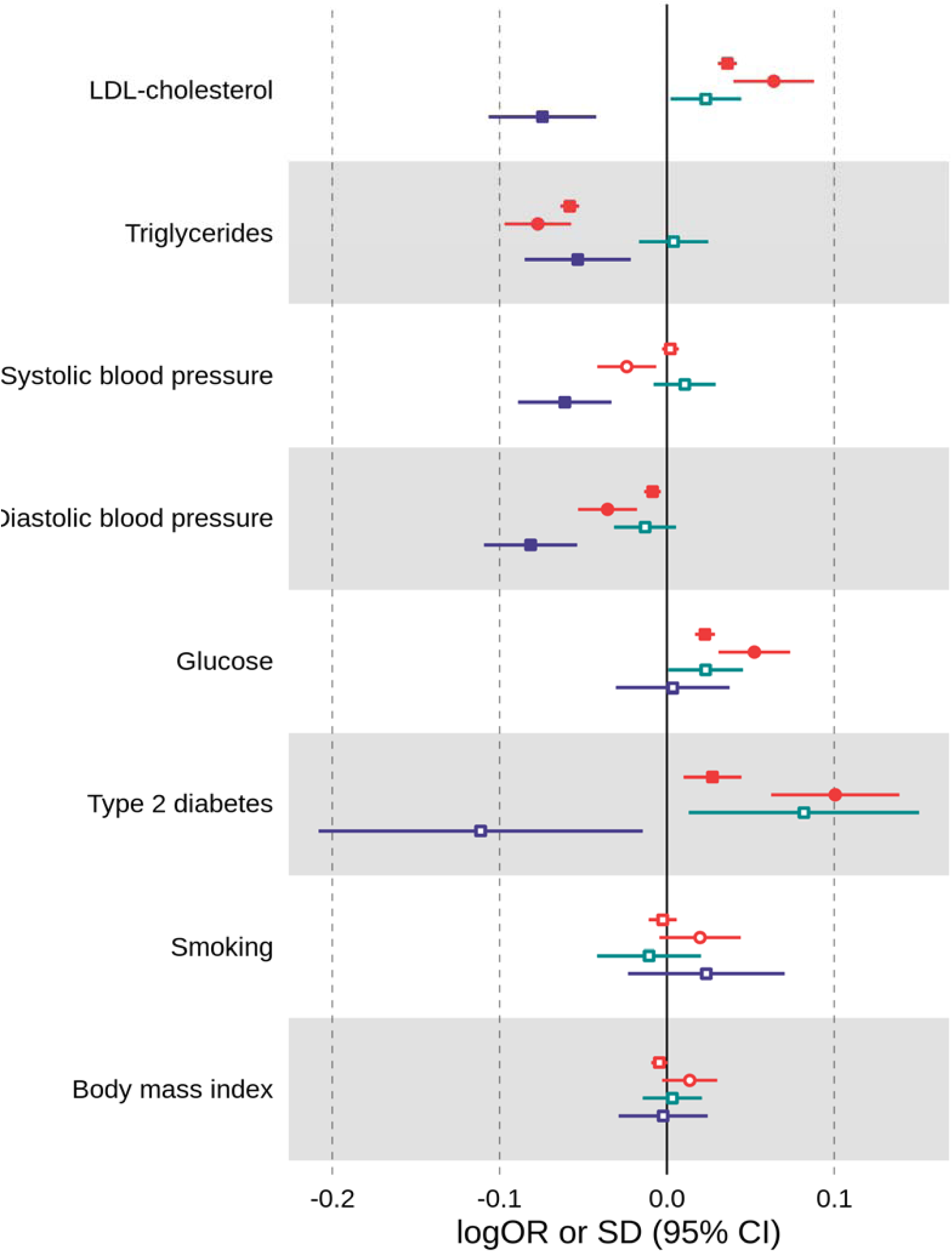
Mendelian randomization results for cardiovascular risk factors related to increasing activity of enzymes coded by *FADS1/2* (D5D/D6D), *ELOVL2* (ELOVL2) and *SCD* (SCD) among individuals of European and East Asian ancestries. Results are expressed as change in standard units or log odds ratio of cardiovascular disease risk factors per standard unit increase in the marker of enzyme activity for *FADS1/2* (i.e. AA:DGLA ratio in Europeans and DGLA:LA ratio in East Asians), *ELOVL2* (i.e. DHA:DPA ratio in Europeans) and *SCD* (i.e. POA:PA ratio in Europeans) loci. For individuals of European ancestry, data was extracted from multiple consortia of genetic association studies. For individuals of East Asian ancestry, data was extracted from BioBank Japan. Full symbols indicate associations at P-value lower than the P-value threshold accounting for multiple testing (P < 0.00625). Smoking is represent by pack years of smoking and number of cigarettes per day in European and East Asian ancestry individuals, respectively. *FADS1/2*: fatty acids desaturase 1/2; *ELOVL2*: elongase 2; *SCD*: stearoyl-CoA desaturase; LDL-cholesterol: low-density lipoprotein-cholesterol.

In the hypothesis-free approach, the *FADS1/2* variant (rs174546) was related not only to fatty acids but also to numerous non-fatty acid traits such as lipid, glycaemic, blood cell traits, physical measures (e.g. pulse, heart rate and height), immune-related disorders (e.g. asthma, hypothyroidism, Crohn’s disease, inflammatory bowel disease) and several biomarkers (e.g. total bilirubin, insulin growth factor (IGF)-1, cystatin C, alkaline phosphatase and urate) among individuals of European ancestry (**Figure 5** and **Supplementary table 4**). The pleiotropic associations of the *FADS1/2* variant (rs174546) were also seen in East Asians in relation to lipid, glycaemic, blood cell traits (**Figure 5** and **Supplementary table 5**). The *ELOVL2* variant (rs3734398) was related to levels of an unknown metabolite X-12627 and DHA and the *SCD* variant (rs603424) was related to multiple SFA/MUFA, as well as to bone mineral density and blood cell-related traits (**Figure 5** and **Supplementary table 4**).

**Figure 5.**
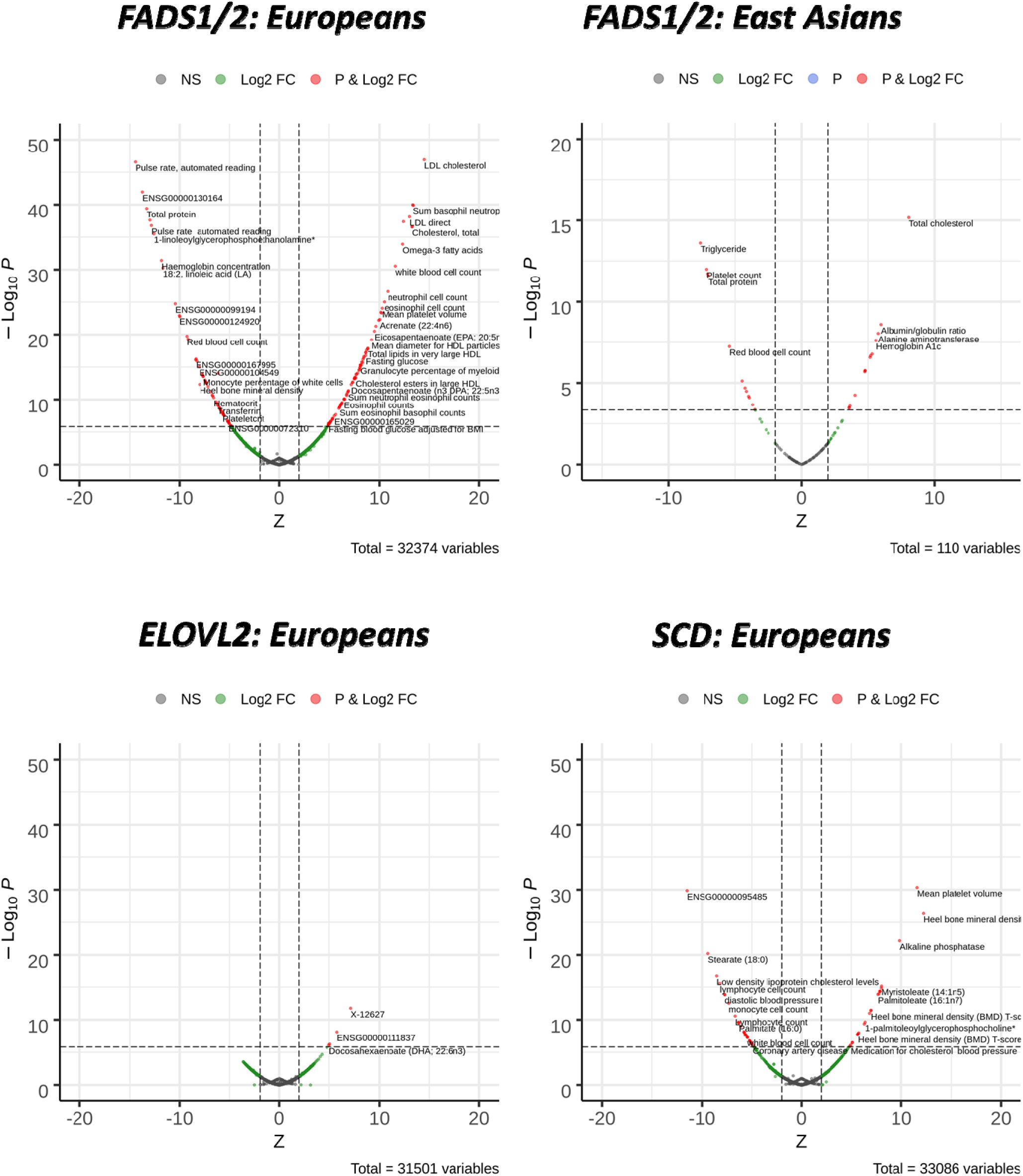
Phenome wide association scan of *FADS1/2* (rs174546), *ELOVL2* (rs3734398) and *SCD* (rs603424) genetic variants in European and East Asian ancestry individuals. Results are expressed as the Z-statistic for the variant-trait association for the allele increasing enzyme expression/activity. Red circles denote P-value < 1.5 x 10^-6^ in Europeans P-value < 4.5 x 10^-4^ in East Asians . *FADS*: fatty acids desaturase; *ELOVL2*: elongase 2; *SCD*: stearoyl-CoA desaturase.

##### Gene expression and tissue-specific analyses

Among Europeans, the selected genetic instruments were strongly associated with the expression of the target genes in one or more tissues (N = 208-670 individuals per tissue). The *FADS1* SNP was related to *FADS1* expression in all tissues analysed, whereas the *ELOVL2* and *SCD* SNPs were related to expression of *ELOVL2* in the liver and expression of *SCD* in (subcutaneous and visceral) adipose tissues, respectively (**Figure 6**). This may be partly related to the expression profile of these genes, since *FADS1* expression is ubiquitous, whereas *ELOVL2* and *SCD* expressions are more prominent in specific tissues (**Supplementary figure 4**). As expected, the alleles increasing the expression of the target genes corresponded to the same alleles increasing the rate of fatty acids biosynthesis, as proxied by markers of enzyme activity, across tissues, except for *FADS1* expression in whole blood (**Figure 6**). Regarding expression of non-target genes, the *FADS1* SNP was related to expression of eleven other genes, including *FADS2*, *FADS3*, *TMEM258*, *MYRF*, and *FEN1*, across multiple tissues, whereas the *ELOVL2* SNP was related to the expression of *SYCP2L* (artery, heart and whole blood tissues) and *ELOVL2-AS1* (liver) and the *SCD* SNP was related to the expression of three other genes, *BLOC1S2*, *PKD2L1* and *OLMALINC*, in subcutaneous adipose tissue only (**Figure 6**).

**Figure 6.**
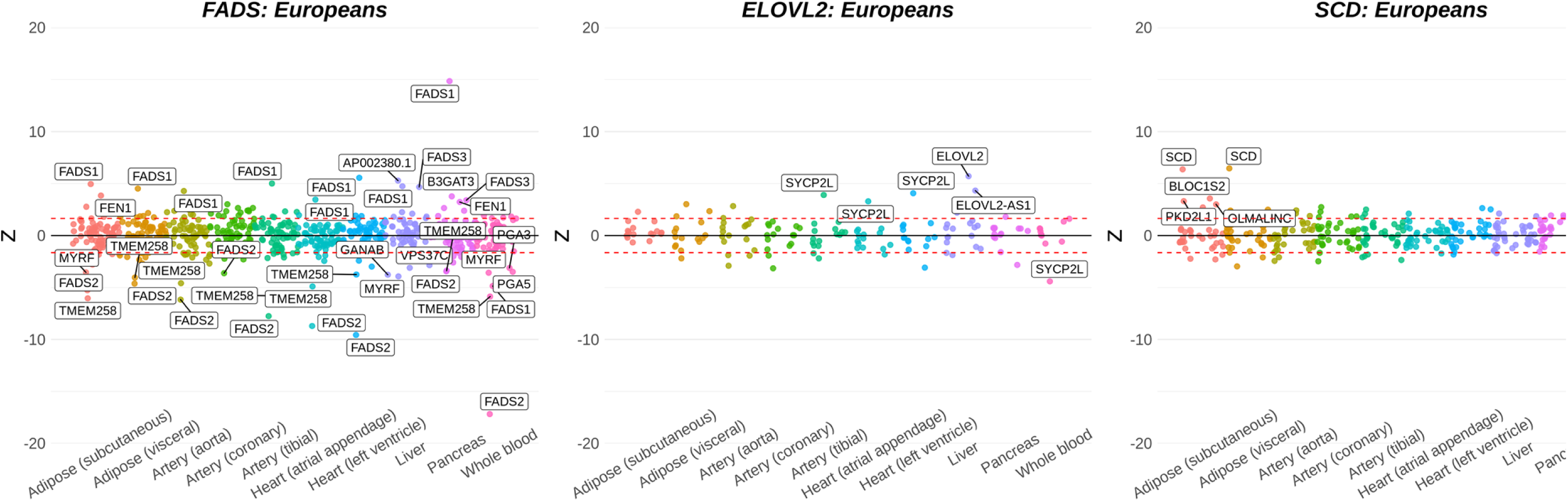
Association of *FADS1/2* (rs174546), *ELOVL2* (rs3734398) and *SCD* (rs603424) genetic variants with expression of cis-genes across multiple tissues among Europeans. Results are expressed as z-statistics (effect estimate / standard error) for the SNP-gene expression association per allele increasing enzyme activity. Only cis-genes (i.e. SNP within 1Mb up- or down-stream of the gene’s transcription start site) were included in the analysis. Genes are labelled in the plot if P-value < 5% FDR per tissue.

The fact that the selected instruments are related to the expression of nearby non-target genes, which is specially the case for the *FADS1* SNP, could bias our analyses if the proteins encoded by these genes directly influence cardiovascular diseases. To explore that, we used multivariable Mendelian randomization, which supported a direct effect of the target genes (i.e. *FADS1*, *ELOVL2* and *SCD*) on cardiovascular diseases and risk factors. In Europeans, higher *FADS1* expression in multiple tissues (e.g. visceral adipose, artery, heart - atrial appendage, liver and pancreas) was related to higher risk of most cardiovascular diseases (**Supplementary figure 5A** and **5B**) and changes in several risk factors, including higher LDL-cholesterol and fasting glucose (**Supplementary figure 6A** and **6B**). Higher expression of *SCD* in subcutaneous adipose tissue was mostly associated to lower risk of coronary artery disease, heart failure, venous thromboembolism and lower diastolic blood pressure, whereas higher expression of *ELOVL2* was related to higher LDL-cholesterol and type 2 diabetes risk (**Supplementary figure 5A-B** and **6A-B**). The conditional F statistics for these analyses ranged from 18 to 433 and 5 to 50 in unadjusted and adjusted models, respectively (**Supplementary table 6**).

#### Confounding by linkage disequilibrium

##### Genetic colocalization

We used genetic colocalization among Europeans to tease apart whether results from Mendelian randomization analyses were compatible with a shared variant between proxies of enzyme activity (i.e. AA:DGLA, DHA:DPAn-3, and POA:PA) and cardiovascular diseases risk and risk factors, which is a necessary (though not sufficient) condition for two traits to be causally related.

For AA:DGLA (proxying D5D activity), there was strong evidence that it colocalised with risk of venous thromboembolism (PPA = 85% for a shared variant). For other cardiovascular disease outcomes, PPA was 0%-27% for a shared variant accompanied by PPA of 60%-100% for the variant being associated with enzyme activity only, which could be a result of limited statistical power (**Supplementary table 7** and **Supplementary figure 7**). In addition, the top variant for AA:DGLA (rs174546) was in strong LD with the top variants for peripheral artery disease, venous thromboembolism, ischemic stroke, and heart failure (R^2^ > 0.8 using 1000G European population), although in weak/moderate LD with top variants for coronary artery disease, atrial fibrillation, and aortic valve stenosis (R^2^ = 0.19- 0.42) (**Supplementary table 8**). After conditioning the genetic association data for AA:DGLA and venous thromboembolism on rs174546, there was no evidence of a distinct or shared genetic signal (**Supplementary table 9**).

There was supportive evidence for a shared causal variant of AA:DGLA with LDL-cholesterol (PPA for shared variant = 87%) and for distinct causal variants with systolic, diastolic blood pressure, and triglycerides (PPA for distinct variants = 90-100%). Evidence was less conclusive for colocalization of AA:DGLA with fasting glucose (PPA for shared variant = 64%) (**Supplementary table 10** and **Supplementary figure 8)**. The top variant for AA:DGLA (rs174546) was in strong LD with the top variants for LDL-cholesterol, triglycerides and glucose (R^2^ > 0.92 using 1000G European population) (**Supplementary table 8**).

For POA:PA (proxying *SCD*/SCD activity), there was strong evidence that it colocalised with risk of coronary artery disease (PPA = 99% for a shared variant) (**Supplementary table 7** and **supplementary figure 7**). After conditioning the genetic association data for POA:PA and coronary artery disease on rs603424, there was no evidence of a distinct or shared genetic signal (**Supplementary table 9**).

Colocalization analyses supported distinct genetic variants between POA:PA and LDL-cholesterol, triglycerides, systolic, diastolic blood pressure, type2 diabetes and body mass index (PPA for distinct variants = 86-100%) (**Supplementary table 10** and **Supplementary figure 8**).

#### Confounding by population stratification, assortative mating and indirect genetic effects

The *FADS1/2* SNP (rs174546) was associated with both negative control outcomes among Europeans: skin colour [mean change of 0.005 unit per C allele (p-value = 5x 10^-2^) and ease of skin tanning [mean change of -0.006 unit per C allele (p-value = 0.007)], while the *SCD* SNP was associated with ease of skin tanning [mean change of -0.006 unit increase per C allele (p-value = 0.037)] (**Supplementary table 11**).

To explore the potential impact of confounding from population stratification, assortative mating and indirect genetic effects, we compared within-sibship associations of the selected genetic variants with cardiovascular risk factors with estimates from unrelated individuals. Estimates were broadly consistent indicating that our findings are unlikely to have been affected by population stratification, assortative mating or indirect genetic effects (**Figure 7**). As an example, for the *FADS1/2* SNP (rs174546), each C allele was related to a mean LDL-cholesterol increase of 0.041 (95% CI: 0.025; 0.057) and 0.036 (95% CI: 0.025; 0.046) SD units within-siblings and in unrelated individuals, respectively.

**Figure 7.**
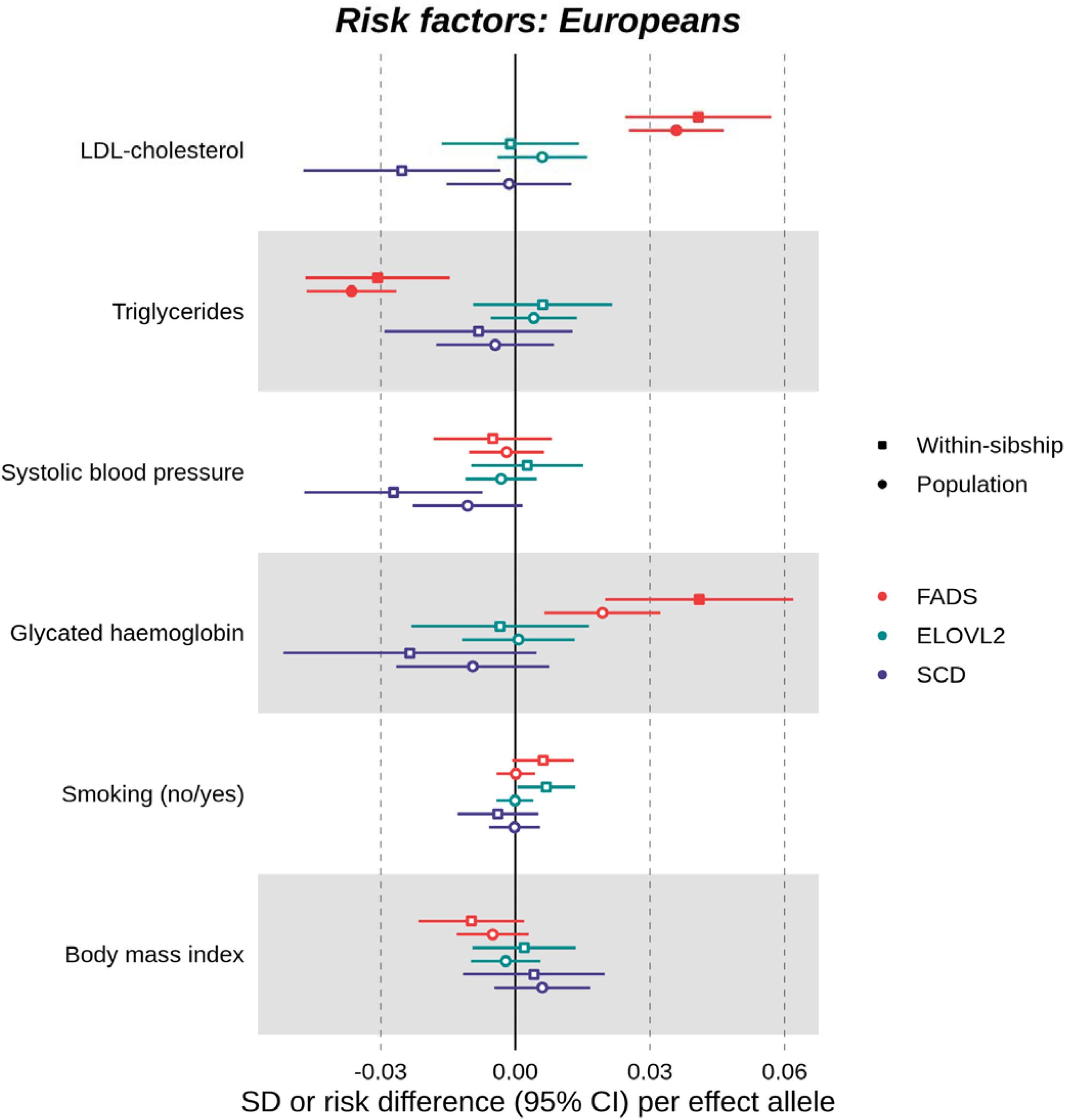
Association of *FADS1/2* (rs174546), *ELOVL2* (rs3734398) and *SCD* (rs603424) genetic variants with cardiovascular risk factors among unrelated individuals and within siblings of European ancestry. Results are expressed as change in standard deviation (SD) units (or risk difference), and 95% confidence intervals (95%CI), of cardiovascular risk factors per allele increasing enzyme activity.

#### Selection bias

##### Positive control analyses

Overall, we observed the expected effect of well-established risk factors on the development of cardiovascular diseases across studies (**Supplementary figure 9**). Systolic, diastolic blood pressure and body mass index were related to higher odds of all cardiovascular disease outcomes in studies of European ancestry individuals (i.e. UK Biobank, genetic association metanalyses and FinnGen) and higher odds of most cardiovascular outcomes (except for coronary and peripheral artery disease) in Biobank Japan. Higher LDL-cholesterol and triglycerides, and liability to type 2 diabetes were related to higher odds of coronary artery disease and peripheral artery disease across studies for both ancestries, while glucose and smoking were related to higher odds of peripheral artery disease in Europeans and East Asians. There were a few instances where these risk factors were related to lower odds of disease, such as type 2 diabetes liability with hemorrhagic stroke

(Biobank Japan) and LDL-cholesterol with hemorrhagic stroke (UK Biobank and Biobank Japan).

## DISCUSSION

### Main findings

In Europeans, our findings indicate that higher PUFA biosynthesis (proxied by *FADS1*/D5D activity) is related to higher risk of several cardiovascular diseases (and risk factors), while higher MUFA biosynthesis (proxied by *SCD*/SCD activity) is related to lower risk of coronary artery disease. In addition, despite the strong LD in the *FADS1/2* region, our results indicate that the relation between PUFA biosynthesis and cardiovascular diseases is driven by changes in *FADS1* (not *FADS2*) expression among Europeans. In East Asians, the same *FADS1/2* variant was related to similar pleiotropic effects on the phenome (e.g. lipid, glycaemic, blood cell traits) compared to Europeans, although the relation with cardiovascular diseases was unclear as most effect estimates were either imprecisely estimated (e.g. atrial fibrillation) or, for coronary artery disease, in the opposite direction in East Asians compared to results in Europeans.

By triangulating multiple approaches, our results are compatible with higher LDL-cholesterol (and possibly glucose) being a downstream effect of higher *FADS1*/D5D activity instead of explained by confounding by LD or by the co-expression of other genes in the region. Given the well-established role of *FADS1*/D5D activity in PUFA biosynthesis and the well-known involvement of LDL-cholesterol in the aetiology of multiple cardiovascular diseases, this strengthens the evidence for a causal relationship and provides a putative mediating pathway for the effect of PUFA biosynthesis on the risk of cardiovascular diseases.

### Previous literature

The relation between fatty acids and cardiovascular diseases has been explored in classical observational studies, randomized controlled trials and Mendelian randomization studies. Most previous studies have focused on coronary artery disease and, to a lesser extent, on stroke; therefore, other types of cardiovascular disease endpoints, such as heart failure and atrial fibrillation, remain under explored

Previous meta-analyses of classical observational studies indicate that higher circulating long-chain omega-3 and omega-6 PUFA are either not associated or are associated with lower risk of coronary artery disease and stroke (68–72), whereas higher circulating MUFA and SFA are either not associated or are associated with higher risk of coronary artery disease and stroke (68, 69). Recent systematic reviews of randomized controlled trials of dietary advice or supplementation of omega-3 and omega-6 PUFA have suggested little to no benefit in reducing the risk of cardiovascular diseases (73–75). However, most studies included in these systematic reviews were at moderate to high risk of bias and there is large uncertainty on the evidence linking PUFA to some cardiovascular outcomes (73–75). It is important to emphasise that comparing our findings to previous classical observational and randomized controlled trials deserves caution as our genetic instruments have a broad impact on the fatty acids pool and, therefore, cannot be used to make inferences about individual fatty acids/fatty acids classes. As an example, higher *FADS1*/D5D activity (instrumented by rs174546) is related to higher longer chain omega-3 and omega-6 PUFA (e.g. AA, EPA and DHA) but lower shorter chain omega-3 and omega-6 PUFA (e.g. LA and ALA).

Several GWAS have reported that SNPs within the *FADS1/2* locus are associated with cardiovascular risk factors (e.g. LDL-cholesterol and triglycerides) (19, 76, 77) and previous Mendelian randomization studies have reported that longer and shorter chain PUFA are related to risk of cardiovascular diseases in contrasting directions among Europeans, including coronary artery disease in CARDIoGRAMplusC4D and UK Biobank (26, 27, 78), ischemic stroke in MEGASTROKE and UK Biobank (28, 29), and venous thromboembolism in UK Biobank (29). Our findings expand on previous Mendelian randomization studies by implicating higher *FADS1*/D5D activity in the development of a wide range of cardiovascular diseases among Europeans in the largest available samples to date (up to 1,153,768 individuals). In addition, to our knowledge, this is the first Mendelian randomization study to report a potential protective effect of higher SCD activity on coronary artery disease among Europeans and to explore the relation between *FADS2*/D6D activity and cardiovascular diseases among East Asians.

### Plausibility of Mendelian randomization assumptions

A major challenge in Mendelian randomization studies is the unprovable assumption that the estimated effect of the genetic instrument on the outcome is mediated by the exposure, and not biased by horizontal pleiotropy, population stratification, assortative mating, indirect genetic effects or selection bias (59, 66, 79). We assessed the plausibility that our findings were explained by these sources of bias through a series of sensitivity analyses.

To mitigate bias due to horizontal pleiotropy (i.e. the genetic instrument influences exposure and outcome via independent pathways), we have restricted our analyses to genetic variants near genes with well-established role in fatty acids biosynthesis. We have confirmed that these variants have the expected impact on the circulating fatty acids pool and on the expression of the target genes in key tissues (except for *FADS1* in whole blood). Previous evidence confirms that the selected *FADS1/2* and *SCD* variants, or variants in high LD, are related to changes in fatty acids composition across multiple sites, including adipose tissue (80–82), brain (83) and liver (84). Genetic colocalization and multivariable Mendelian randomization (adjusting for co-expressed genes in the region) supported a causal relation between D5D activity, venous thromboembolism and LDL-cholesterol, and between SCD activity and coronary artery disease. It is important to note that we were likely underpowered to test colocalization between fatty acids biosynthesis and cardiovascular disease outcomes. Where there was evidence that the selected genetic variant was associated with the expression of non-target genes in the region in a given tissue, findings from multivariable Mendelian randomization were consistent with expression of the target gene (i.e. *FADS1*) having direct effects on the outcome.

Confounding could be introduced in Mendelian randomization studies due to population stratification, assortative mating and indirect genetic effects. Of these factors, population stratification is likely to be of the most concern for this study (59). Despite attempts to control for population structure in genetic association data, there could still be residual population structure (63). We showed that within-sibship associations of *FADS1/2*, *ELOVL2*, and *SCD* variants with established cardiovascular risk factors were broadly similar to estimates from unrelated individuals, suggesting that our results are unlikely to be affected by population stratification, assortative mating or indirect genetic effects of parents.

Non-random sample selection may introduce bias in Mendelian randomization studies especially if the mechanism of selection depends on the exposure and/or outcome (66, 85). Using a positive control approach, we were able to identify the expected effect of well-established risk factors on cardiovascular diseases across studies contributing with data on cardiovascular disease endpoints, which is reassuring given our concerns that case-control ascertainment could introduce bias in the analyses. Although results from the positive control approach argues against selection being a major source of bias in this study, we cannot fully rule out that selection might have introduced some bias in our analyses as bias due to selection will depend on context-specific causal structures underlying the data under consideration.

### Implications

Our findings are supportive of the involvement of fatty acids biosynthesis in the aetiology of cardiovascular diseases and highlight D5D and SCD as potential targets for the prevention of these diseases. Further work is needed to understand the precise underlying mechanism(s) are required.

The relation between D5D activity and cardiovascular diseases is plausibly mediated by one or more fatty acids involved in the PUFA biosynthesis pathway. Given the ubiquitous impact of higher D5D activity on the circulating PUFA pool, we cannot pinpoint which specific fatty acids are driving these effects. For illustration, higher D5D activity decreases LA and ALA (and other omega-3 and omega-6 PUFA upstream of the reaction catalysed by D5D). Lower LA may relate to unfavourable metabolic changes, such as higher plasma LDL-cholesterol, apolipoprotein B, and triglycerides, and haemoglobin A1c (2, 86) and, therefore, is a plausible mediator of the relation between higher D5D activity and higher cardiovascular diseases risk. On the other hand, higher D5D activity increases long-chain PUFA such as AA, which influences key membrane/tissue functions, such as membrane fluidity, the activity of membrane-bound receptors, transport proteins and signal transmission (87), and is a precursor for eicosanoids (e.g. prostaglandins, leukotrienes, and thromboxane), which are involved in inflammation, platelet aggregation and vascular remodeling (88).

The putative mechanisms underpinning the relation SCD activity and coronary artery disease in humans are unclear. *Scd-1* deficient rodents are protected against diet-induced obesity, insulin resistance, and hepatic steatosis (89–91), but show increased inflammation and atherogenesis (91, 92). The putative protective effect of higher SCD activity on coronary artery disease might be related to lower availability of palmitic acid and consequent lower production of its toxic metabolites, such as ceramides (93).

### Conclusions

We found supportive evidence for an involvement of PUFA and MUFA biosynthesis in the aetiology of cardiovascular diseases. Our study illustrates the power of integrating multiple approaches to improve causal inference on the role of modifiable risk factors in the development of cardiovascular diseases.

## Data Availability

Genetic association data on fatty acids, cardiovascular diseases (CVDs) and CVDs risk factors are publicly available or available on request from original investigators as detailed in Supplementary tables 1 and 2.

http://www.cardiogramplusc4d.org/data-downloads/

http://www.megastroke.org/

http://kp4cd.org/datasets/mi

http://csg.sph.umich.edu/willer/public/afib2018/

http://jenger.riken.jp/en/result

https://www.finngen.fi/en/access_results

## Supplemental information description

Supplementary methods
Supplementary tables 1 to 10
Supplementary figures 1 to 9

## Acknowledgements

We want to acknowledge participants and investigators from FinnGen study, UK Biobank, Biobank Japan, and the multiple large-scale GWAS consortia which made summary data available, including CARDIoGRAMplusC4D, MEGASTROKE, HERMES, atrial fibrillation GWAS metanalysis, aortic aneurysm GWAS metanalysis, CHARGE, and the Within Family Consortium. This work was carried out using UK Biobank project 15825. Data on coronary artery disease have been contributed by CARDIoGRAMplusC4D investigators and have been downloaded from www.CARDIOGRAMPLUSC4D.ORG’. The MEGASTROKE project received funding from sources specified at http://www.megastroke.org/acknowledgments.html (<u>a</u> list of all <u>MEGASTROKE authors</u> can be found in the appendix A - Supplementary methods). This work was carried out using the computational facilities of the Advanced Computing Research Centre, University of Bristol - http://www.bristol.ac.uk/acrc/. Quality Control filtering of the UK Biobank data was conducted by R.Mitchell, G.Hemani, T.Dudding, L.Corbin, S.Harrison, L.Paternoster as described in the published protocol (doi: 10.5523/bris.1ovaau5sxunp2cv8rcy88688v). The MRC IEU UK Biobank GWAS pipeline was developed by B.Elsworth, R.Mitchell, C.Raistrick, L.Paternoster, G.Hemani, T.Gaunt (doi: 10.5523/bris.pnoat8cxo0u52p6ynfaekeigi).

## Web resources

Coloc: https://cran.r-project.org/web/packages/coloc/index.html

GWIS: https://sites.google.com/site/mgnivard/gwis

IEU Open GWAS Project: https://gwas.mrcieu.ac.uk/

MVMR R package: https://github.com/WSpiller/MVMR

PWCoCco: https://github.com/jwr-git/pwcoco

TwoSampleMR R package: https://mrcieu.github.io/TwoSampleMR

