## Supplementary methods for "The impact of fatty acids biosynthesis on the risk of cardiovascular diseases in Europeans and East Asians: *A Mendelian randomization study*"

*Data sources for genetic associations with cardiovascular disease outcomes*

UK Biobank is a population-based cohort of approximately 500,000 (5.5% of those invited) people aged 40 to 69 years when recruited during 2006-2010 from several centres across the United Kingdom (1, 2). Details on study design, participants and quality control methods have been described previously (3, 4). UK Biobank received ethical approval from the Research Ethics Committee (REC reference for UK Biobank is 11/NW/0382). This work was carried out using UK Biobank project 15825.

FinnGen is a public-private partnership project combining genotype data from Finnish biobanks and digital health record data from Finnish health registries (https://www.finngen.fi/en). Six regional and three country-wide Finnish biobanks participate in FinnGen (i.e. Auria Biobank, Biobank Borealis of Northern Finland, Biobank of Eastern Finland, Central Finland Biobank, Finnish Red Cross Blood Service Biobank, Finnish Clinical Biobank Tampere, Helsinki Biobank, Terveystalo Biobank, and THL Biobank). We used FinnGen data from release 4, which includes 176,899 participants (5).

BioBank Japan project is a registry-based study of approximately 200,000 individuals recruited from 2003 to 2008 (mean age 62.7 years for men and 61.5 years for women) enrolled at 12 cooperative medical institutes in Japan. Details on study design, participants and quality control methods have been described previously (6-9). The study protocol was approved by the research ethics committees at the Institute of Medical Science, the University of Tokyo, the RIKEN Yokohama Institute, and the 12 cooperating hospitals. All participants gave written consent to participate in the study.

The procedures used to generate genetic association data for cardiovascular outcomes have been described previously for UK Biobank (4), FinnGen (5), and BioBank Japan (8, 9).

Large-scale genetic consortia of cardiovascular disease outcomes in studies targeting individuals of European ancestry only, or predominantly, included the *Coronary Artery Disease Genome-Wide Replication and Meta-analysis Plus the Coronary Artery Disease Genetics Consortium* (CARDIoGRAMplusC4D) (N = 184,305) (10), MEGASTROKE (N = 446,696) (11), *The Heart Failure Molecular Epidemiology for Therapeutic Targets* (HERMES) (N = 977,323) (12), an atrial fibrillation genetic association metanalysis (N = 1,030,836) (13), and an abdominal aortic aneurysm genetic association metanalysis (N = 117,970) (14).

*Genetic effect on enzyme activity*

Enzyme activity was proxied based on enzyme-specific product to substrate ratio as follows:

| Locus | Chr | Enzyme | Activity proxy | Fatty acids class | Ancestry |
| --- | --- | --- | --- | --- | --- |
| *FADS1* | 11 | D5D | AA:DGLA | PUFA n-6 | Europeans  East Asians |
| *FADS2* | 11 | D6D | GLA:LA  DGLA:LA | PUFA n-6 | Europeans  East Asians |
| *ELOVL2* | 6 | ELOVL2 | DHA:DPAn3 | PUFA n-3 | Europeans |
| *SCD* | 10 | SCD | POA:PA | MUFA/SFA | Europeans |

AA: arachidonic acid; DGLA: dihomo-γ-linolenic acid; DHA: docosahexaenoic acid; DPA: docosapentaenoic acid; *ELOVL2*: elongase 2; *FADS*: fatty acids desaturase; GLA: γ-linoleic acid; LA: linoleic acid; MUFA: monounsaturated fatty acids; PA: palmitic acid; POA: palmitoleic acid; PUFA: polyunsaturated fatty acids; SFA: saturated fatty acids; *SCD*: stearoyl-CoA desaturase.

For Europeans, we derived genetic association data for enzyme activity proxies using GWIS (“Genome-wide Inferred Study”). GWIS is a method that provides an approximation of GWAS summary statistics for a variable that is a function of phenotypes for which GWAS summary statistics, phenotypic means, and covariances are available. SNP-trait effect estimates are derived as a linear function of the allele frequencies, population means of measured traits (assumed to approximate the intercepts of the model) and SNP-trait effect estimates of measured traits. Corresponding standard errors can be derived using the Delta-method having obtained the covariance matrix for effect estimates (15).

For East Asians, genetic association data for D5D, proxied by AA:DGLA, and D6D , proxied by DGLA:LA, has been previously generated by the original GWAS investigations and, therefore, we used the original data rather than approximating the rations using GWIS.

*Mendelian randomization analysis*

For each cardiovascular outcome, we used the Wald ratio method to estimate the odds ratio of disease for each standard unit increase in the proxy of enzyme activity:

$$\beta_{MR}= \frac{\beta_{y}}{\beta_{x}}$$

and corresponding standard error:

$$\sigma_{MR}= \frac{\sigma_{y}}{\beta_{x}}$$

Where $\beta_{y}$ and $\beta_{x}$ are the coefficients for the association of the genetic variant with the outcome (Y) and the exposure (X), respectively, and $\sigma_{y}$ is the standard error for the association of the genetic variant with Y.

*Approximating R^2^ and F-statistics from genetic association data with fatty acids traits*

For each selected SNP, we approximated the R^2^, a measure of the variance in exposure explained by the genetic variant, and the F-statistics, a measure of instrument strength (16), as follows:

$$R^{2}=2*\beta_{gx}^{2}*MAF*(1-MAF)$$

$$F=\left( \frac{n-k-1}{k} \right)*(\frac{R^{2}}{1-R^{2}})$$

where:

$\beta_{gx}$ = SNP-fatty acid trait association estimate (in standard deviation units)

MAF = minor allele frequency

n = sample size

k = number of SNPs

*Exploring bias in Mendelian randomization analyses*

Genetic colocalization

*Coloc* enumerates all possible configurations of causal variants for each of two traits (e.g. fatty acid- and cardiovascular disease-related traits), and uses a Bayesian approach to calculate support for each causal model (H_0_: no association; H_1_: association with trait 1 only; H_2_: association with trait 2 only; H_3_: association with both traits due to distinct causal variants; H_4_: association with both traits due to a single shared causal variant~~s~~). We restricted *coloc* analysis to a genomic region within a 500-kb window around each target gene (*FADS1/2*, *ELOVL2*, and *SCD*) and assumed prior probabilities that any random SNP in the region is associated with trait 1 (p1 = $1\times{10}^{-4}$), trait 2 (p2 = $1\times{10}^{-4}$), or both traits (p12 = $1\times{10}^{-6}$).

Gene expression and tissue-specific analyses

We explored the influence of higher expression of the target genes (i.e. *FADS1/2*, *ELOVL2* and *SCD*), and their tissue specificity, on cardiovascular disease risk and risk factors by integrating expression quantitative trait loci (eQTL) data with genetic association data for cardiovascular traits. These analyses consisted of three steps:

i. Performing a cross-tissue assessment of the association of the genetic variants selected for the main analyses with transcription of any genes in the region (aka cis-genes) (i.e. SNP within 1Mb up- or down-stream of the gene’s transcription start site). Cis-genes were selected for follow-up analysis if the P-value for the SNP-gene expression association was below the 5% false discovery rate (FDR) threshold for each tissue.
ii. Selecting independent eQTLs (P < $5\times{10}^{-5}$; R^2^ < 0.05) for each target gene (i.e. *FADS1/2*, *ELOVL2*, and *SCD*) to test the effect of target gene expression on cardiovascular outcomes in a two-sample Mendelian randomization framework. LD across variants (R^2^) was estimated using 1000G reference populations for Europeans (i.e. CEU) and East Asians (i.e. EAS).
iii. Given some eQTLs might influence or be in LD with eQTLs influencing the expression of other (non-target) genes, we used multivariable Mendelian randomization to test the independent effect of higher expression of the target gene (adjusting for co-expression of other cis-genes) on cardiovascular outcomes across tissues using the MVMR R package (17). This analysis allowed us to estimate the direct contribution of changes in the expression of each target gene even when eQTLs were related to co-expression of non- target genes. For the third step, eQTLs were selected as described in second step with an additional round of LD clumping (R^2^ < 0.05) to ensure only independent SNPs were included in the model as independent eQTLs select for a gene could be in LD with eQTLs selected for a different gene.

Multivariable Mendelian randomization models were estimated for each combination of target gene, co-expressed gene, outcome and tissue if (a) the target gene SNP (i.e. rs174546, rs2236212, and rs603424) was related to expression of non- target genes in that tissue (step i), (b) more than two independent SNPs were selected for the analyses, and (c) the conditional F statistics for the target gene expression was equal or higher than 5.

For Europeans, we extracted eQTL data from the GTEx project v8 for multiple tissues of relevance to cardiovascular diseases — i.e. subcutaneous/visceral adipose tissues, aorta/coronary/tibial arteries, heart, liver, pancreas and whole blood (N = 208-670 individuals per tissue) (18). For east Asians, we extracted eQTL data from Biobank Japan for whole blood and blood cells (N = 98-105 individuals per tissue) given data was not available for the tissues of interest (19).

**Appendix A** – List of investigators from the MEGASTROKE CONSORTIUM

Rainer Malik ^1^, Ganesh Chauhan ^2^, Matthew Traylor ^3^, Muralidharan Sargurupremraj ^4,5^, Yukinori Okada ^6,7,8^, Aniket Mishra ^4,5^, Loes Rutten-Jacobs ^3^, Anne-Katrin Giese ^9^, Sander W van der Laan ^10^, Solveig Gretarsdottir ^11^, Christopher D Anderson ^12,13,14,14^, Michael Chong ^15^, Hieab HH Adams ^16,17^, Tetsuro Ago ^18^, Peter Almgren ^19^, Philippe Amouyel ^20,21^, Hakan Ay ^22,13^, Traci M Bartz ^23^, Oscar R Benavente ^24^, Steve Bevan ^25^, Giorgio B Boncoraglio ^26^, Robert D Brown, Jr.  ^27^, Adam S Butterworth ^28,29^, Caty Carrera ^30,31^, Cara L Carty ^32,33^, Daniel I Chasman ^34,35^, Wei-Min Chen ^36^, John W Cole ^37^, Adolfo Correa ^38^, Ioana Cotlarciuc ^39^, Carlos Cruchaga ^40,41^, John Danesh ^28,42,43,44^, Paul IW de Bakker ^45,46^, Anita L DeStefano ^47,48^, Marcel den Hoed ^49^, Qing Duan ^50^, Stefan T Engelter ^51,52^, Guido J Falcone ^53,54^, Rebecca F Gottesman ^55^, Raji P Grewal ^56^, Vilmundur Gudnason ^57,58^, Stefan Gustafsson ^59^, Jeffrey Haessler ^60^, Tamara B Harris ^61^, Ahamad Hassan ^62^, Aki S Havulinna ^63,64^, Susan R Heckbert ^65^, Elizabeth G Holliday ^66,67^, George Howard ^68^, Fang-Chi Hsu ^69^, Hyacinth I Hyacinth ^70^, M Arfan Ikram ^16^, Erik Ingelsson ^71,72^, Marguerite R Irvin ^73^, Xueqiu Jian ^74^, Jordi Jiménez-Conde ^75^, Julie A Johnson ^76,77^, J Wouter Jukema ^78^, Masahiro Kanai ^6,7,79^, Keith L Keene ^80,81^, Brett M Kissela ^82^, Dawn O Kleindorfer ^82^, Charles Kooperberg ^60^, Michiaki Kubo ^83^, Leslie A Lange ^84^, Carl D Langefeld ^85^, Claudia Langenberg ^86^, Lenore J Launer ^87^, Jin-Moo Lee ^88^, Robin Lemmens ^89,90^, Didier Leys ^91^, Cathryn M Lewis ^92,93^, Wei-Yu Lin ^28,94^, Arne G Lindgren ^95,96^, Erik Lorentzen ^97^, Patrik K Magnusson ^98^, Jane Maguire ^99^, Ani Manichaikul ^36^, Patrick F McArdle ^100^, James F Meschia ^101^, Braxton D Mitchell ^100,102^, Thomas H Mosley ^103,104^, Michael A Nalls ^105,106^, Toshiharu Ninomiya ^107^, Martin J O'Donnell ^15,108^, Bruce M Psaty ^109,110,111,112^, Sara L Pulit ^113,45^, Kristiina Rannikmäe ^114,115^, Alexander P Reiner ^65,116^, Kathryn M Rexrode ^117^, Kenneth Rice ^118^, Stephen S Rich ^36^, Paul M Ridker ^34,35^, Natalia S Rost ^9,13^, Peter M Rothwell ^119^, Jerome I Rotter ^120,121^, Tatjana Rundek ^122^, Ralph L Sacco ^122^, Saori Sakaue ^7,123^, Michele M Sale ^124^, Veikko Salomaa ^63^, Bishwa R Sapkota ^125^, Reinhold Schmidt ^126^, Carsten O Schmidt  ^127^, Ulf Schminke ^128^, Pankaj Sharma ^39^, Agnieszka Slowik ^129^, Cathie LM Sudlow ^114,115^, Christian Tanislav ^130^, Turgut Tatlisumak ^131,132^, Kent D Taylor ^120,121^, Vincent NS Thijs ^133,134^, Gudmar Thorleifsson ^11^, Unnur Thorsteinsdottir ^11^, Steffen Tiedt ^1^, Stella Trompet ^135^, Christophe Tzourio ^5,136,137^, Cornelia M van Duijn ^138,139^, Matthew Walters ^140^, Nicholas J Wareham ^86^, Sylvia Wassertheil-Smoller ^141^, James G Wilson ^142^, Kerri L Wiggins ^109^, Qiong Yang ^47^, Salim Yusuf ^15^, Najaf Amin ^16^, Hugo S Aparicio ^185,48^, Donna K Arnett ^186^, John Attia ^187^, Alexa S Beiser ^47,48^, Claudine Berr ^188^, Julie E Buring ^34,35^, Mariana Bustamante ^189^, Valeria Caso ^190^, Yu-Ching Cheng ^191^, Seung Hoan Choi ^192,48^, Ayesha Chowhan ^185,48^, Natalia Cullell ^31^, Jean-François Dartigues ^193,194^, Hossein Delavaran ^95,96^, Pilar Delgado ^195^, Marcus Dörr ^196,197^, Gunnar Engström ^19^, Ian Ford ^198^, Wander S Gurpreet ^199^, Anders Hamsten ^200,201^, Laura Heitsch ^202^, Atsushi Hozawa ^203^, Laura Ibanez ^204^, Andreea Ilinca ^95,96^, Martin Ingelsson ^205^, Motoki Iwasaki ^206^, Rebecca D Jackson ^207^, Katarina Jood ^208^, Pekka Jousilahti ^63^, Sara Kaffashian ^4,5^, Lalit Kalra ^209^, Masahiro Kamouchi ^210^, Takanari Kitazono ^211^, Olafur Kjartansson ^212^, Manja Kloss ^213^, Peter J Koudstaal ^214^, Jerzy Krupinski ^215^, Daniel L Labovitz ^216^, Cathy C Laurie ^118^, Christopher R Levi ^217^, Linxin Li ^218^, Lars Lind ^219^, Cecilia M Lindgren ^220,221^, Vasileios Lioutas ^222,48^, Yong Mei Liu ^223^, Oscar L Lopez ^224^, Hirata Makoto ^225^, Nicolas Martinez-Majander ^172^, Koichi Matsuda ^225^, Naoko Minegishi ^203^, Joan Montaner  ^226^, Andrew P Morris ^227,228^, Elena Muiño ^31^, Martina Müller-Nurasyid ^229,230,231^, Bo Norrving ^95,96^, Soichi Ogishima ^203^, Eugenio A Parati ^232^, Leema Reddy Peddareddygari ^56^, Nancy L Pedersen ^98,233^, Joanna Pera ^129^, Markus Perola ^63,234^, Alessandro Pezzini ^235^, Silvana Pileggi ^236^, Raquel Rabionet ^237^, Iolanda Riba-Llena ^30^, Marta Ribasés ^238^, Jose R Romero ^185,48^, Jaume Roquer ^239,240^, Anthony G Rudd ^241,242^, Antti-Pekka Sarin ^243,244^, Ralhan Sarju ^199^, Chloe Sarnowski ^47,48^, Makoto Sasaki ^245^, Claudia L Satizabal ^185,48^, Mamoru Satoh ^245^, Naveed Sattar ^246^, Norie Sawada ^206^, Gerli Sibolt ^172^, Ásgeir Sigurdsson ^247^, Albert Smith ^248^, Kenji Sobue ^245^, Carolina Soriano-Tárraga ^240^, Tara Stanne ^249^, O Colin Stine ^250^, David J Stott ^251^, Konstantin Strauch ^229,252^, Takako Takai  ^203^, Hideo Tanaka ^253,254^, Kozo Tanno ^245^, Alexander Teumer ^255^, Liisa Tomppo ^172^, Nuria P Torres-Aguila ^31^, Emmanuel Touze ^256,257^, Shoichiro Tsugane  ^206^, Andre G Uitterlinden ^258^, Einar M Valdimarsson ^259^, Sven J van der Lee ^16^, Henry Völzke ^255^, Kenji Wakai  ^253^, David Weir ^260^, Stephen R Williams ^261^, Charles DA Wolfe ^241,242^, Quenna Wong ^118^, Huichun Xu ^191^, Taiki Yamaji ^206^, Dharambir K Sanghera ^125,169,170^, Olle Melander ^19^, Christina Jern ^171^, Daniel Strbian ^172,173^, Israel Fernandez-Cadenas ^31,30^, W T Longstreth, Jr ^174,65^, Arndt Rolfs ^175^, Jun Hata ^107^, Daniel Woo ^82^, Jonathan Rosand ^12,13,14^, Guillaume Pare ^15^, Jemma C Hopewell ^176^, Danish Saleheen ^177^, Kari Stefansson ^11,178^, Bradford B Worrall ^179^, Steven J Kittner ^37^, Sudha Seshadri ^180,48^, Myriam Fornage ^74,181^, Hugh S Markus ^3^, Joanna MM Howson ^28^, Yoichiro Kamatani ^6,182^, Stephanie Debette ^4,5^, Martin Dichgans ^1,183,184^

1 Institute for Stroke and Dementia Research (ISD), University Hospital, LMU Munich, Munich, Germany

2 Centre for Brain Research, Indian Institute of Science, Bangalore, India

3 Stroke Research Group, Division of Clinical Neurosciences, University of Cambridge, UK

4 INSERM U1219 Bordeaux Population Health Research Center, Bordeaux, France

5 University of Bordeaux, Bordeaux, France

6 Laboratory for Statistical Analysis, RIKEN Center for Integrative Medical Sciences, Yokohama, Japan

7 Department of Statistical Genetics, Osaka University Graduate School of Medicine, Osaka, Japan

8 Laboratory of Statistical Immunology, Immunology Frontier Research Center (WPI-IFReC), Osaka University, Suita, Japan.

9 Department of Neurology, Massachusetts General Hospital, Harvard Medical School, Boston, MA, USA

10 Laboratory of Experimental Cardiology, Division of Heart and Lungs, University Medical Center Utrecht, University of Utrecht, Utrecht,Netherlands

11 deCODE genetics/AMGEN inc, Reykjavik, Iceland

12 Center for Genomic Medicine, Massachusetts General Hospital (MGH), Boston, MA, USA

13 J. Philip Kistler Stroke Research Center, Department of Neurology, MGH, Boston, MA, USA

14 Program in Medical and Population Genetics, Broad Institute, Cambridge, MA, USA

15 Population Health Research Institute, McMaster University, Hamilton, Canada

16 Department of Epidemiology, Erasmus University Medical Center, Rotterdam, Netherlands

17 Department of Radiology and Nuclear Medicine, Erasmus University Medical Center, Rotterdam, Netherlands

18 Department of Medicine and Clinical Science, Graduate School of Medical Sciences, Kyushu University, Fukuoka, Japan

19 Department of Clinical Sciences, Lund University, Malmö, Sweden

20 Univ. Lille, Inserm, Institut Pasteur de Lille, LabEx DISTALZ-UMR1167, Risk factors and molecular determinants of aging-related diseases, F-59000 Lille, France

21 Centre Hosp. Univ Lille, Epidemiology and Public Health Department, F-59000 Lille, France

22 AA Martinos Center for Biomedical Imaging, Department of Radiology, Massachusetts General Hospital, Harvard Medical School, Boston, MA, USA

23 Cardiovascular Health Research Unit, Departments of Biostatistics and Medicine, University of Washington, Seattle, WA, USA

24 Division of Neurology, Faculty of Medicine, Brain Research Center, University of British Columbia, Vancouver, Canada

25 School of Life Science, University of Lincoln, Lincoln, UK

26 Department of Cerebrovascular Diseases, Fondazione IRCCS Istituto Neurologico "Carlo Besta", Milano, Italy

27 Department of Neurology, Mayo Clinic Rochester, Rochester, MN, USA

28 MRC/BHF Cardiovascular Epidemiology Unit, Department of Public Health and Primary Care, University of Cambridge, Cambridge, UK

29 The National Institute for Health Research Blood and Transplant Research Unit in Donor Health and Genomics, University of Cambridge, UK

30 Neurovascular Research Laboratory, Vall d'Hebron Institut of Research, Neurology and Medicine Departments-Universitat Autònoma de Barcelona, Vall d’Hebrón Hospital, Barcelona, Spain

31 Stroke Pharmacogenomics and Genetics, Fundacio Docència i Recerca MutuaTerrassa, Terrassa, Spain

32 Children's Research Institute, Children's National Medical Center, Washington, DC, USA

33 Center for Translational Science, George Washington University, Washington, DC, USA

34 Division of Preventive Medicine, Brigham and Women's Hospital, Boston, MA, USA

35 Harvard Medical School, Boston, MA, USA

36 Center for Public Health Genomics, Department of Public Health Sciences, University of Virginia, Charlottesville, VA, USA

37 Department of Neurology, University of Maryland School of Medicine and Baltimore VAMC, Baltimore, MD, USA

38 Departments of Medicine, Pediatrics and Population Health Science, University of Mississippi Medical Center, Jackson, MS, USA

39 Institute of Cardiovascular Research, Royal Holloway University of London, UK  &  Ashford and St Peters Hospital, Surrey UK

40 Department of Psychiatry,The Hope Center Program on Protein Aggregation and Neurodegeneration (HPAN),Washington University, School of Medicine, St. Louis, MO, USA

41 Department of Developmental Biology, Washington University School of Medicine, St. Louis, MO, USA

42 NIHR Blood and Transplant Research Unit in Donor Health and Genomics, Department of Public Health and Primary Care, University of Cambridge, Cambridge, UK

43 Wellcome Trust Sanger Institute, Wellcome Trust Genome Campus, Hinxton,  Cambridge, UK

44 British Heart Foundation, Cambridge Centre of Excellence, Department of Medicine, University of Cambridge, Cambridge, UK

45 Department of Medical Genetics, University Medical Center Utrecht, Utrecht, Netherlands

46 Department of Epidemiology, Julius Center for Health Sciences and Primary Care, University Medical Center Utrecht, Utrecht, Netherlands

47 Boston University School of Public Health, Boston, MA, USA

48 Framingham Heart Study, Framingham, MA, USA

49 Department of Immunology, Genetics and Pathology and Science for Life Laboratory, Uppsala University, Uppsala, Sweden

50 Department of Genetics, University of North Carolina, Chapel Hill, NC, USA

51 Department of Neurology and Stroke Center, Basel University Hospital, Switzerland

52 Neurorehabilitation Unit, University and University Center for Medicine of Aging and Rehabilitation Basel, Felix Platter Hospital, Basel, Switzerland

53 Department of Neurology, Yale University School of Medicine, New Haven, CT, USA

54 Program in Medical and Population Genetics, The Broad Institute of Harvard and MIT, Cambridge, MA, USA

55 Department of Neurology, Johns Hopkins University School of Medicine, Baltimore, MD, USA

56 Neuroscience Institute, SF Medical Center, Trenton, NJ, USA

57 Icelandic Heart Association Research Institute, Kopavogur, Iceland

58 University of Iceland, Faculty of Medicine, Reykjavik, Iceland

59 Department of Medical Sciences, Molecular Epidemiology and Science for Life Laboratory, Uppsala University, Uppsala, Sweden

60 Division of Public Health Sciences, Fred Hutchinson Cancer Research Center, Seattle, WA, USA

61 Laboratory of Epidemiology and Population Science, National Institute on Aging, National Institutes of Health, Bethesda, MD, USA

62 Department of Neurology, Leeds General Infirmary, Leeds Teaching Hospitals NHS Trust, Leeds, UK

63 National Institute for Health and Welfare, Helsinki, Finland

64 FIMM - Institute for Molecular Medicine Finland, Helsinki, Finland

65 Department of Epidemiology, University of Washington, Seattle, WA, USA

66 Public Health Stream, Hunter Medical Research Institute, New Lambton, Australia

67 Faculty of Health and Medicine, University of Newcastle, Newcastle, Australia

68 School of Public Health, University of Alabama at Birmingham, Birmingham, AL, USA

69 Department of Biostatistical Sciences, Wake Forest School of Medicine, Winston-Salem, NC, USA

70 Aflac Cancer and Blood Disorder Center, Department of Pediatrics, Emory University School of Medicine, Atlanta, GA, USA

71 Department of Medicine, Division of Cardiovascular Medicine, Stanford University School of Medicine, CA, USA

72 Department of Medical Sciences, Molecular Epidemiology and Science for Life Laboratory, Uppsala University, Uppsala, Sweden

73 Epidemiology, School of Public Health, University of Alabama at Birmingham, USA

74 Brown Foundation Institute of Molecular Medicine, University of Texas Health Science Center at Houston, Houston, TX, USA

75 Neurovascular Research Group (NEUVAS), Neurology Department, Institut Hospital del Mar d'Investigació Mèdica, Universitat Autònoma de Barcelona, Barcelona, Spain

76 Department of Pharmacotherapy and Translational Research and Center for Pharmacogenomics, University of Florida, College of Pharmacy, Gainesville, FL, USA

77 Division of Cardiovascular Medicine, College of Medicine, University of Florida, Gainesville, FL, USA

78 Department of Cardiology, Leiden University Medical Center, Leiden, the Netherlands

79 Program in Bioinformatics and Integrative Genomics, Harvard Medical School, Boston, MA, USA

80 Department of Biology, East Carolina University, Greenville, NC, USA

81 Center for Health Disparities, East Carolina University, Greenville, NC, USA

82 University of Cincinnati College of Medicine, Cincinnati, OH, USA

83 RIKEN Center for Integrative Medical Sciences, Yokohama, Japan

84 Department of Medicine, University of Colorado Denver, Anschutz Medical Campus, Aurora, CO, USA

85 Center for Public Health Genomics and Department of Biostatistical Sciences, Wake Forest School of Medicine, Winston-Salem, NC, USA

86 MRC Epidemiology Unit, University of Cambridge School of Clinical Medicine, Institute of Metabolic Science, Cambridge Biomedical Campus, Cambridge, UK

87 Intramural Research Program, National Institute on Aging, National Institutes of Health, Bethesda, MD, USA

88 Department of Neurology, Radiology, and Biomedical Engineering, Washington University School of Medicine, St. Louis, MO, USA

89 KU Leuven – University of Leuven, Department of Neurosciences,  Experimental Neurology, Leuven, Belgium

90 VIB Center for Brain & Disease Research, University Hospitals Leuven, Department of Neurology, Leuven, Belgium

91 Univ.-Lille, INSERM U 1171. CHU Lille. Lille, France

92 Department of Medical and Molecular Genetics, King's College London, London, UK

93 SGDP Centre, Institute of Psychiatry, Psychology & Neuroscience, King's College London, London, UK

94 Northern Institute for Cancer Research, Paul O'Gorman Building, Newcastle University, Newcastle, UK

95 Department of Clinical Sciences Lund, Neurology, Lund University, Lund, Sweden

96 Department of Neurology and Rehabilitation Medicine, Skåne University Hospital, Lund, Sweden

97 Bioinformatics Core Facility, University of Gothenburg, Gothenburg, Sweden

98 Department of Medical Epidemiology and Biostatistics, Karolinska Institutet, Stockholm, Sweden

99 University of Technology Sydney, Faculty of Health, Ultimo, Australia

100 Department of Medicine, University of Maryland School of Medicine, MD, USA

101 Department of Neurology, Mayo Clinic, Jacksonville, FL, USA

102 Geriatrics Research and Education Clinical Center, Baltimore Veterans Administration Medical Center, Baltimore, MD, USA

103 Division of Geriatrics, School of Medicine, University of Mississippi Medical Center, Jackson, MS, USA

104 Memory Impairment and Neurodegenerative Dementia Center, University of Mississippi Medical Center, Jackson, MS, USA

105 Laboratory of Neurogenetics, National Institute on Aging, National institutes of Health, Bethesda, MD, USA

106 Data Tecnica International, Glen Echo MD, USA

107 Department of Epidemiology and Public Health, Graduate School of Medical Sciences, Kyushu University, Fukuoka, Japan

108 Clinical Research Facility, Department of Medicine, NUI Galway, Galway, Ireland

109 Cardiovascular Health Research Unit, Department of Medicine, University of Washington, Seattle, WA, USA

110 Department of Epidemiology, University of Washington, Seattle, WA

111 Department of Health Services, University of Washington, Seattle, WA, USA

112 Kaiser Permanente Washington Health Research Institute, Seattle, WA, USA

113 Brain Center Rudolf Magnus, Department of Neurology, University Medical Center Utrecht, Utrecht, The Netherlands

114 Usher Institute of Population Health Sciences and Informatics, University of Edinburgh, Edinburgh, UK

115 Centre for Clinical Brain Sciences, University of Edinburgh, Edinburgh, UK

116 Fred Hutchinson Cancer Research Center, University of Washington, Seattle, WA, USA

117 Department of Medicine, Brigham and Women's Hospital, Boston, MA, USA

118 Department of Biostatistics, University of Washington, Seattle, WA, USA

119 Nuffield Department of Clinical Neurosciences, University of Oxford, UK

120 Institute for Translational Genomics and Population Sciences, Los Angeles Biomedical Research Institute at  Harbor-UCLA Medical Center, Torrance, CA, USA

121 Division of Genomic Outcomes, Department of Pediatrics, Harbor-UCLA Medical Center, Torrance, CA, USA

122 Department of Neurology, Miller School of Medicine, University of Miami, Miami, FL, USA

123 Department of Allergy and Rheumatology, Graduate School of Medicine, the University of Tokyo, Tokyo, Japan

124 Center for Public Health Genomics, University of Virginia, Charlottesville, VA, USA

125 Department of Pediatrics, College of Medicine, University of Oklahoma Health Sciences Center, Oklahoma City, OK, USA

126 Department of Neurology, Medical University of Graz, Graz, Austria

127 University Medicine  Greifswald, Institute for Community Medicine, SHIP-KEF, Greifswald, Germany

128 University Medicine  Greifswald,  Department of Neurology, Greifswald, Germany

129 Department of Neurology, Jagiellonian University, Krakow, Poland

130 Department of Neurology, Justus Liebig University, Giessen, Germany

131 Department of Clinical Neurosciences/Neurology, Institute of Neuroscience and Physiology, Sahlgrenska Academy at University of Gothenburg, Gothenburg, Sweden

132 Sahlgrenska University Hospital, Gothenburg, Sweden

133 Stroke Division, Florey Institute of Neuroscience and Mental Health, University of Melbourne, Heidelberg, Australia

134 Austin Health, Department of Neurology, Heidelberg, Australia

135 Department of Internal Medicine, Section Gerontology and Geriatrics, Leiden University Medical Center, Leiden, the Netherlands

136 INSERM U1219, Bordeaux, France

137 Department of Public Health, Bordeaux University Hospital, Bordeaux, France

138 Genetic Epidemiology Unit, Department of Epidemiology, Erasmus University Medical Center Rotterdam, Netherlands

139 Center for Medical Systems Biology, Leiden, Netherlands

140 School of Medicine, Dentistry and Nursing at the University of Glasgow, Glasgow, UK

141 Department of Epidemiology and Population Health, Albert Einstein College of Medicine, NY, USA

142 Department of Physiology and Biophysics, University of Mississippi Medical Center, Jackson, MS, USA

143 A full list of members and affiliations appears in the Supplementary Note

144 Department of Human Genetics, McGill University, Montreal, Canada

145 Department of Pathophysiology, Institute of Biomedicine and Translation Medicine, University of Tartu, Tartu, Estonia

146 Department of Cardiac Surgery, Tartu University Hospital, Tartu, Estonia

147 Clinical Gene Networks AB,Stockholm, Sweden

148 Department of Genetics and Genomic Sciences, The Icahn Institute for Genomics and Multiscale Biology Icahn School of Medicine at Mount Sinai, New York, NY , USA

149 Department of Pathophysiology, Institute of Biomedicine and Translation Medicine, University of Tartu, Biomeedikum, Tartu, Estonia

150 Integrated Cardio Metabolic Centre, Department of Medicine, Karolinska Institutet, Karolinska Universitetssjukhuset, Huddinge, Sweden.

151 Clinical Gene Networks AB, Stockholm, Sweden

152 Sorbonne Universités, UPMC Univ. Paris 06, INSERM, UMR_S 1166, Team Genomics & Pathophysiology of Cardiovascular Diseases, Paris, France

153 ICAN Institute for Cardiometabolism and Nutrition, Paris, France

154 Department of Biomedical Engineering, University of Virginia, Charlottesville, VA, USA

155 Group Health Research Institute, Group Health Cooperative, Seattle, WA, USA

156 Seattle Epidemiologic Research and Information Center, VA Office of Research and Development, Seattle, WA, USA

157 Cardiovascular Research Center, Massachusetts General Hospital, Boston, MA, USA

158 Department of Medical Research, Bærum Hospital, Vestre Viken Hospital Trust, Gjettum, Norway

159 Saw Swee Hock School of Public Health, National University of Singapore and National University Health System, Singapore

160 National Heart and Lung Institute, Imperial College London, London, UK

161 Department of Gene Diagnostics and Therapeutics, Research Institute, National Center for Global Health and Medicine, Tokyo, Japan

162 Department of Epidemiology, Tulane University School of Public Health and Tropical Medicine, New Orleans, LA, USA

163 Department of Cardiology,University Medical Center Groningen, University of Groningen, Netherlands

164 MRC-PHE Centre for Environment and Health, School of Public Health, Department of Epidemiology and Biostatistics, Imperial College London, London, UK

165 Department of Epidemiology and Biostatistics, Imperial College London, London, UK

166 Department of Cardiology, Ealing Hospital NHS Trust, Southall, UK

167 National Heart, Lung and Blood Research Institute, Division of Intramural Research, Population Sciences Branch, Framingham, MA, USA

168 A full list of members and affiliations appears at the end of the manuscript

169 Department of Phamaceutical Sciences, Collge of Pharmacy, University of Oklahoma Health Sciences Center, Oklahoma City, OK, USA

170 Oklahoma Center for Neuroscience, Oklahoma City, OK, USA

171 Department of Pathology and Genetics, Institute of Biomedicine, The Sahlgrenska Academy at University of Gothenburg, Gothenburg, Sweden

172 Department of Neurology, Helsinki University Hospital, Helsinki, Finland

173 Clinical Neurosciences, Neurology, University of Helsinki, Helsinki, Finland

174 Department of Neurology, University of Washington, Seattle, WA, USA

175 Albrecht Kossel Institute, University Clinic of Rostock, Rostock, Germany

176 Clinical Trial Service Unit and Epidemiological Studies Unit, Nuffield Department of Population Health, University of Oxford, Oxford, UK

177 Department of Genetics, Perelman School of Medicine, University of Pennsylvania, PA, USA

178 Faculty of Medicine, University of Iceland, Reykjavik, Iceland

179 Departments of Neurology and Public Health Sciences, University of Virginia School of Medicine, Charlottesville, VA, USA

180 Department of Neurology, Boston University School of Medicine, Boston, MA, USA

181 Human Genetics Center, University of Texas Health Science Center at Houston, Houston, TX, USA

182 Center for Genomic Medicine, Kyoto University Graduate School of Medicine, Kyoto, Japan

183 Munich Cluster for Systems Neurology (SyNergy), Munich, Germany

184 German Center for Neurodegenerative Diseases (DZNE), Munich, Germany

185 Boston University School of Medicine, Boston, MA, USA

186 University of Kentucky College of Public Health, Lexington, KY, USA

187 University of Newcastle and Hunter Medical Research Institute, New Lambton, Australia

188 Univ. Montpellier, Inserm, U1061, Montpellier, France

189 Centre for Research in Environmental Epidemiology, Barcelona, Spain

190 Department of Neurology, Università degli Studi di Perugia, Umbria, Italy

191 Department of Medicine, University of Maryland School of Medicine, Baltimore, MD, USA

192 Broad Institute, Cambridge, MA, USA

193 Univ. Bordeaux, Inserm, Bordeaux Population Health Research Center, UMR 1219, Bordeaux, France

194 Bordeaux University Hospital, Department of Neurology, Memory Clinic, Bordeaux, France

195 Neurovascular Research Laboratory. Vall d'Hebron Institut of Research, Neurology and Medicine Departments-Universitat Autònoma de Barcelona. Vall d’Hebrón Hospital, Barcelona, Spain

196 University Medicine Greifswald, Department of Internal Medicine B, Greifswald, Germany

197 DZHK, Greifswald, Germany

198 Robertson Center for Biostatistics, University of Glasgow, Glasgow, UK

199 Hero DMC Heart Institute, Dayanand Medical College & Hospital, Ludhiana, India

200 Atherosclerosis Research Unit, Department of Medicine Solna, Karolinska Institutet, Stockholm, Sweden

201 Karolinska Institutet, Stockholm, Sweden

202 Division of Emergency Medicine, and Department of Neurology, Washington University School of Medicine, St. Louis, MO, USA

203 Tohoku Medical Megabank Organization, Sendai, Japan

204 Department of Psychiatry, Washington University School of Medicine, St. Louis, MO, USA

205 Department of Public Health and Caring Sciences / Geriatrics, Uppsala University, Uppsala, Sweden

206 Epidemiology and Prevention Group, Center for Public Health Sciences, National Cancer Center, Tokyo, Japan

207 Department of Internal Medicine and the Center for Clinical and Translational Science, The Ohio State University, Columbus, OH, USA

208 Institute of Neuroscience and Physiology, the Sahlgrenska Academy at University of Gothenburg, Goteborg, Sweden

209 Department of Basic and Clinical Neurosciences, King's College London, London, UK

210 Department of Health Care Administration and Management, Graduate School of Medical Sciences, Kyushu University, Japan

211 Department of Medicine and Clinical Science, Graduate School of Medical Sciences, Kyushu University, Japan

212 Landspitali National University Hospital, Departments of Neurology & Radiology, Reykjavik, Iceland

213 Department of Neurology, Heidelberg University Hospital, Germany

214 Department of Neurology, Erasmus University Medical Center

215 Hospital Universitari Mutua Terrassa, Terrassa (Barcelona), Spain

216 Albert Einstein College of Medicine, Montefiore Medical Center, New York, NY, USA

217 John Hunter Hospital, Hunter Medical Research Institute and University of Newcastle, Newcastle, NSW, Australia

218 Centre for Prevention of Stroke and Dementia, Nuffield Department of Clinical Neurosciences, University of Oxford, UK

219 Department of Medical Sciences, Uppsala University, Uppsala, Sweden

220 Genetic and Genomic Epidemiology Unit, Wellcome Trust Centre for Human Genetics, University of Oxford, Oxford, UK

221 The Wellcome Trust Centre for Human Genetics, Oxford, UK

222 Beth Israel Deaconess Medical Center, Boston, MA, USA

223 Wake Forest School of Medicine, Wake Forest, NC, USA

224 Department of Neurology, University of Pittsburgh, Pittsburgh, PA, USA

225 BioBank Japan, Laboratory of Clinical Sequencing, Department of Computational biology and medical Sciences, Graduate school of Frontier Sciences, The University of Tokyo, Tokyo, Japan

226 Neurovascular Research Laboratory, Vall d'Hebron Institut of Research, Neurology and Medicine Departments-Universitat Autònoma de Barcelona. Vall d’Hebrón Hospital, Barcelona, Spain

227 Department of Biostatistics, University of Liverpool, Liverpool, UK

228 Wellcome Trust Centre for Human Genetics, University of Oxford, Oxford, UK

229 Institute of Genetic Epidemiology, Helmholtz Zentrum München - German Research Center for Environmental Health, Neuherberg, Germany

230 Department of Medicine I, Ludwig-Maximilians-Universität, Munich, Germany

231 DZHK (German Centre for Cardiovascular Research), partner site Munich Heart Alliance, Munich, Germany

232 Department of Cerebrovascular Diseases, Fondazione IRCCS Istituto Neurologico “Carlo Besta”, Milano, Italy

233 Karolinska Institutet, MEB, Stockholm, Sweden

234 University of Tartu, Estonian Genome Center, Tartu, Estonia, Tartu, Estonia

235 Department of Clinical and Experimental Sciences, Neurology Clinic, University of Brescia, Italy

236 Translational Genomics Unit, Department of Oncology, IRCCS Istituto di Ricerche Farmacologiche Mario Negri, Milano, Italy

237 Department of Genetics, Microbiology and Statistics, University of Barcelona, Barcelona, Spain

238 Psychiatric Genetics Unit, Group of Psychiatry, Mental Health and Addictions, Vall d’Hebron Research Institute (VHIR), Universitat Autònoma de Barcelona, Biomedical Network Research Centre on Mental Health (CIBERSAM), Barcelona, Spain

239 Department of Neurology, IMIM-Hospital del Mar, and Universitat Autònoma de Barcelona, Spain

240 IMIM (Hospital del Mar Medical Research Institute), Barcelona, Spain

241 National Institute for Health Research Comprehensive Biomedical Research Centre, Guy's & St. Thomas' NHS Foundation Trust and King's College London, London, UK

242 Division of Health and Social Care Research, King's College London, London, UK

243 FIMM-Institute for Molecular Medicine Finland, Helsinki, Finland

244 THL-National Institute for Health and Welfare, Helsinki, Finland

245 Iwate Tohoku Medical Megabank Organization, Iwate Medical University, Iwate, Japan

246 BHF Glasgow Cardiovascular Research Centre, Faculty of Medicine, Glasgow, UK

247 deCODE Genetics/Amgen, Inc., Reykjavik, Iceland

248 Icelandic Heart Association, Reykjavik, Iceland

249 Institute of Biomedicine, the Sahlgrenska Academy at University of Gothenburg, Goteborg, Sweden

250 Department of Epidemiology, University of Maryland School of Medicine, Baltimore, MD, USA

251 Institute of Cardiovascular and Medical Sciences, Faculty of Medicine, University of Glasgow, Glasgow, UK

252 Chair of Genetic Epidemiology, IBE, Faculty of Medicine, LMU Munich, Germany

253 Division of Epidemiology and Prevention, Aichi Cancer Center Research Institute, Nagoya, Japan

254 Department of Epidemiology, Nagoya University Graduate School of Medicine, Nagoya, Japan

255 University Medicine Greifswald, Institute for Community Medicine, SHIP-KEF, Greifswald, Germany

256 Department of Neurology, Caen University Hospital, Caen, France

257 University of Caen Normandy, Caen, France

258 Department of Internal Medicine, Erasmus University Medical Center, Rotterdam, Netherlands

259 Landspitali University Hospital, Reykjavik, Iceland

260 Survey Research Center, University of Michigan, Ann Arbor, MI, USA

261 University of Virginia Department of Neurology, Charlottesville, VA, USA
