## Supplementary figures for "The impact of fatty acids biosynthesis on the risk of cardiovascular diseases in Europeans and East Asians: *A Mendelian randomization study*"

**Supplementary figure 1**. Effect of *FADS*, *ELOVL2* and *SCD* genetic variants on circulating fatty acids in the discovery and replication datasets among individuals of European ancestry.

Results are expressed as z-statistics (effect estimate / standard error) for the SNP-fatty acids association. Blue, red, and grey boxes denote, respectively, decreases, increases, and no change in fatty acids per allele increasing enzyme activity, while white boxes represent missing data. Asterisks indicate P value: < $5\times{10}^{-8}$ (***), < $5\times{10}^{-5}$ (**), and < $5\times{10}^{-2}$ (*).

Plasma FA: plasma fatty acids; N: median sample size used for estimating SNP-fatty acids association; *FADS*: fatty acids desaturase; *ELOVL2*: elongase 2; *SCD*: stearoyl-CoA desaturase; PUFA: polyunsaturated fatty acids; ALA: α-linolenic acid; SDA: stearidonic acid; ETA: eicosatetraenoic acid; EPA: eicosapentaenoic acid; DPA n-3: docosapentaenoic acid; TPA n-3: tetracosapentaenoic acid; DHA: docosahexaenoic acid; LA: linoleic acid; GLA: γ-linolenic acid; EDA: eicosadienoic acid; DGLA: dihomo-γ-linoleic acid; AA: arachidonic acid; ADA: adrenic acid; DPA n-6: docosapentaenoic acid; MUFA: monounsaturated fatty acids; POA: palmitoleic acid; OA: oleic acid; SFA: saturated fatty acids; PA: palmitic acid; SA: stearic acid.

**Supplementary figure 2**. Effect of *FADS* genetic variant on circulating fatty acids among individuals of East Asian ancestry.

Results are expressed as z-statistics (effect estimate / standard error) for the SNP-fatty acids association. Blue, red, and grey boxes denote, respectively, decreases, increases, and no change in fatty acids per allele increasing enzyme activity. Asterisks indicate P value: < $5\times{10}^{-8}$ (***), < $5\times{10}^{-5}$ (**), and < $5\times{10}^{-2}$ (*).

Plasma FA: plasma fatty acids; N: sample size used for estimating SNP-fatty acids association; PUFA: polyunsaturated fatty acids; ALA: α-linolenic acid; EPA: eicosapentaenoic acid; DHA: docosahexaenoic acid; LA: linoleic acid; GLA: γ-linolenic acid; DGLA: dihomo-γ-linoleic acid; AA: arachidonic acid.

**Supplementary figure 3**. Mendelian randomization results for the risk of cardiovascular diseases related to increasing activity of enzymes coded by *FADS1/2* (D5D/D6D), *ELOVL2* (ELOVL2) and *SCD* (SCD) among individuals of European ancestry.

Results are expressed as odds ratio of cardiovascular diseases per standard unit increase in the marker of enzyme activity for *FADS* (i.e. AA:DGLA ratio), *ELOVL2* (i.e. DHA:DPA n-3 ratio) and *SCD* (i.e. POA:PA ratio) loci. SNP-cardiovascular diseases association data were extracted from multiple GWAS not including UK Biobank (“GWAS”), UK Biobank (“UKB”), GWAS including UK Biobank (“GWAS+UKB”), FinnGen (“FinnGen”). Full symbols indicate associations at P-value lower than the P-value threshold accounting multiple testing among Europeans (P < 0.006).

AA: arachidonic acid; DGLA: dihomo-γ-linoleic acid; DHA: docosahexaenoic acid; DPA: docosapentaenoic acid; LA: linoleic acid; PA: palmitic acid; POA: palmitoleic acid; SNP: single nucleotide polymorphism; *FADS*: fatty acids desaturase; *ELOVL2*: elongase 2; *SCD*: stearoyl-CoA desaturase.

**Supplementary figure 4**. Expression of target genes across multiple tissues. Plots extracted from GTEx portal [<https://www.gtexportal.org/home>].

**Supplementary figure 5A**. Effect of higher expression of target genes (*FADS1*, *ELOVL2*, *SCD*) across tissues on the risk of cardiovascular diseases (i.e. coronary artery disease, ischemic stroke, hemorrhagic stroke and heart failure)

Results are expressed as log odds ratio of cardiovascular diseases per standard unit increase in gene expression unadjusted (univariable Mendelian randomization) or adjusted for co-expressed cis-genes (multivariable Mendelian randomization). Target genes: *FADS1*: fatty acids desaturase 1; *ELOVL2*: elongase 2; *SCD*: stearoyl-CoA desaturase. Co-expressed cis-genes: *BLOC1S2*: biogenesis of lysosomal organelles complex 1 subunit 2; *FADS2*: fatty acids desaturase 2; *TMEM258*: transmembrane protein 258; *MYRF*: myelin regulatory factor; *PKD2L1*: polycystin 2 like 1, transient receptor potential cation channel.

**Supplementary figure 5B**. Effect of higher expression of target genes (*FADS1*, *ELOVL2*, *SCD*) across tissues on the risk of cardiovascular diseases (i.e. atrial fibrillation, peripheral artery disease, aortic aneurysm, venous thromboembolism and aortic valve stenosis)

Results are expressed as log odds ratio of cardiovascular diseases per standard unit increase in gene expression unadjusted (univariable Mendelian randomization) or adjusted for co-expressed cis-genes (multivariable Mendelian randomization). Target genes: *FADS1*: fatty acids desaturase 1; *ELOVL2*: elongase 2; *SCD*: stearoyl-CoA desaturase. Co-expressed cis-genes: *BLOC1S2*: biogenesis of lysosomal organelles complex 1 subunit 2; *FADS2*: fatty acids desaturase 2; *TMEM258*: transmembrane protein 258; *MYRF*: myelin regulatory factor; *PKD2L1*: polycystin 2 like 1, transient receptor potential cation channel.

**Supplementary figure 6A**. Effect of higher expression of target genes (*FADS1*, *ELOVL2*, *SCD*) across tissues on cardiovascular risk factors (i.e. LDL-cholesterol, triglycerides, diastolic and systolic blood pressure)

Results are expressed as changes in standard units of risk factors per standard unit increase in gene expression unadjusted (univariable Mendelian randomization) or adjusted for co-expressed cis-genes (multivariable Mendelian randomization). Target genes: *FADS1*: fatty acids desaturase 1; *ELOVL2*: elongase 2; *SCD*: stearoyl-CoA desaturase. Co-expressed cis-genes: *BLOC1S2*: biogenesis of lysosomal organelles complex 1 subunit 2; *FADS2*: fatty acids desaturase 2; *TMEM258*: transmembrane protein 258; *MYRF*: myelin regulatory factor; *PKD2L1*: polycystin 2 like 1, transient receptor potential cation channel.

**Supplementary figure 6B**. Effect of higher expression of target genes (*FADS1*, *ELOVL2*, *SCD*) across tissues on cardiovascular risk factors (i.e. fasting glucose, type 2 diabetes, body mass index and smoking)

Results are expressed as changes in standard units of risk factors per standard unit increase in gene expression unadjusted (univariable Mendelian randomization) or adjusted for co-expressed cis-genes (multivariable Mendelian randomization). Target genes: *FADS1*: fatty acids desaturase 1; *ELOVL2*: elongase 2; *SCD*: stearoyl-CoA desaturase. Co-expressed cis-genes: *BLOC1S2*: biogenesis of lysosomal organelles complex 1 subunit 2; *FADS2*: fatty acids desaturase 2; *TMEM258*: transmembrane protein 258; *MYRF*: myelin regulatory factor; *PKD2L1*: polycystin 2 like 1, transient receptor potential cation channel.

**Supplementary figure 7**. Stacked association plots of fatty acids enzyme activity (proxied by AA:DGLA, DHA:DPA, and POA:PA ratio) on cardiovascular diseases risk among individuals of European ancestry (pooled data).

Results for each trait are expressed as log_10_ P-values for the *FADS, ELOVL2*, and *SCD* locus (columns 1, 2, and 3, respectively). AA: arachidonic acid; DGLA: dihomo-γ-linoleic acid; DHA: docosahexaenoic acid; DPA: docosapentaenoic acid; LA: linoleic acid; PA: palmitic acid; POA: palmitoleic acid; *FADS*: fatty acids desaturase; *ELOVL2*: elongase 2; *SCD*: stearoyl-CoA desaturase; CAD: coronary artery disease; AIS: any ischemic stroke; AHS: any hemorrhagic stroke; HF: heart failure; AF: atrial fibrillation; PAD: peripheral artery disease; AA: aortic aneurysm; VT: venous thromboembolism; AVS: aortic valve stenosis.

**Supplementary figure 8**. Stacked association plots of fatty acids enzyme activity (proxied by AA:DGLA, DHA:DPA, and POA:PA ratio) on cardiovascular risk factors among individuals of European ancestry.

Results for each trait are expressed as log_10_ P-values for the *FADS, ELOVL2*, and *SCD* locus (columns 1, 2, and 3, respectively). AA: arachidonic acid; DGLA: dihomo-γ-linoleic acid; DHA: docosahexaenoic acid; DPA: docosapentaenoic acid; LA: linoleic acid; PA: palmitic acid; POA: palmitoleic acid; *FADS*: fatty acids desaturase; *ELOVL2*: elongase 2; *SCD*: stearoyl-CoA desaturase; LDLc: low-density lipoprotein-cholesterol; TG: triglycerides; SBP: systolic blood pressure; DBP: diastolic blood pressure; Glc: fasting glucose; T2D: type 2 diabetes; SMK: smoking; BMI: body mass index.

**Supplementary figure 9**. Effect of well-established risk factors on the development of cardiovascular diseases using genetic association cardiovascular disease data from UK Biobank (UKB), genetic association metanalyses (GWAS), FinnGen (FinnGen) and Biobank Japan (BBJ) participants.

Results are expressed as log odds ratio of disease per unit increase in risk factors (i.e. continuous risk factors in standard deviation units and type 2 diabetes in log odds). The inverse variance weighted estimator, a standard two-sample Mendelian randomization method, was used to estimate the effect (and corresponding 95% confidence intervals) of risk factors on the odds of cardiovascular diseases. Smoking is represented as pack years of smoking in results for Europeans and cigarettes per day in results for East Asians.

**Supplementary figure 1**

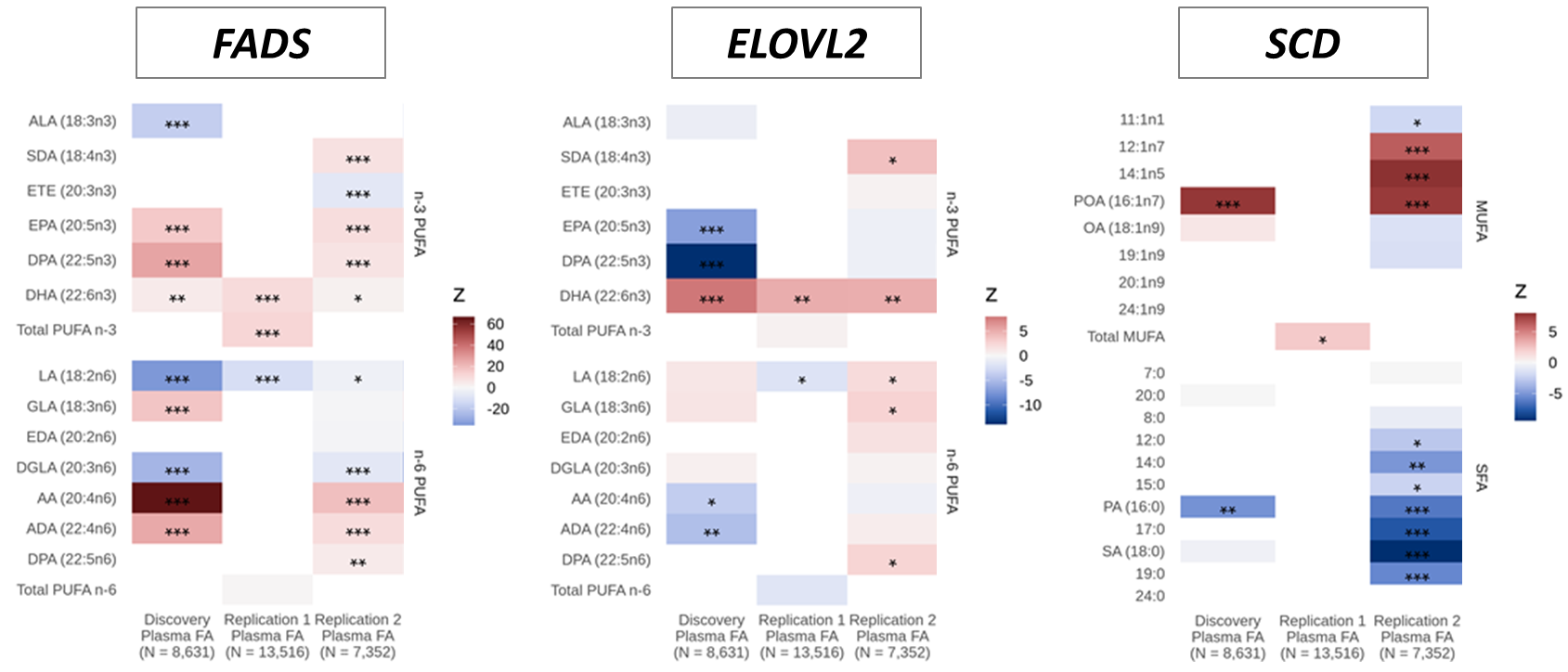

**Supplementary figure 2**

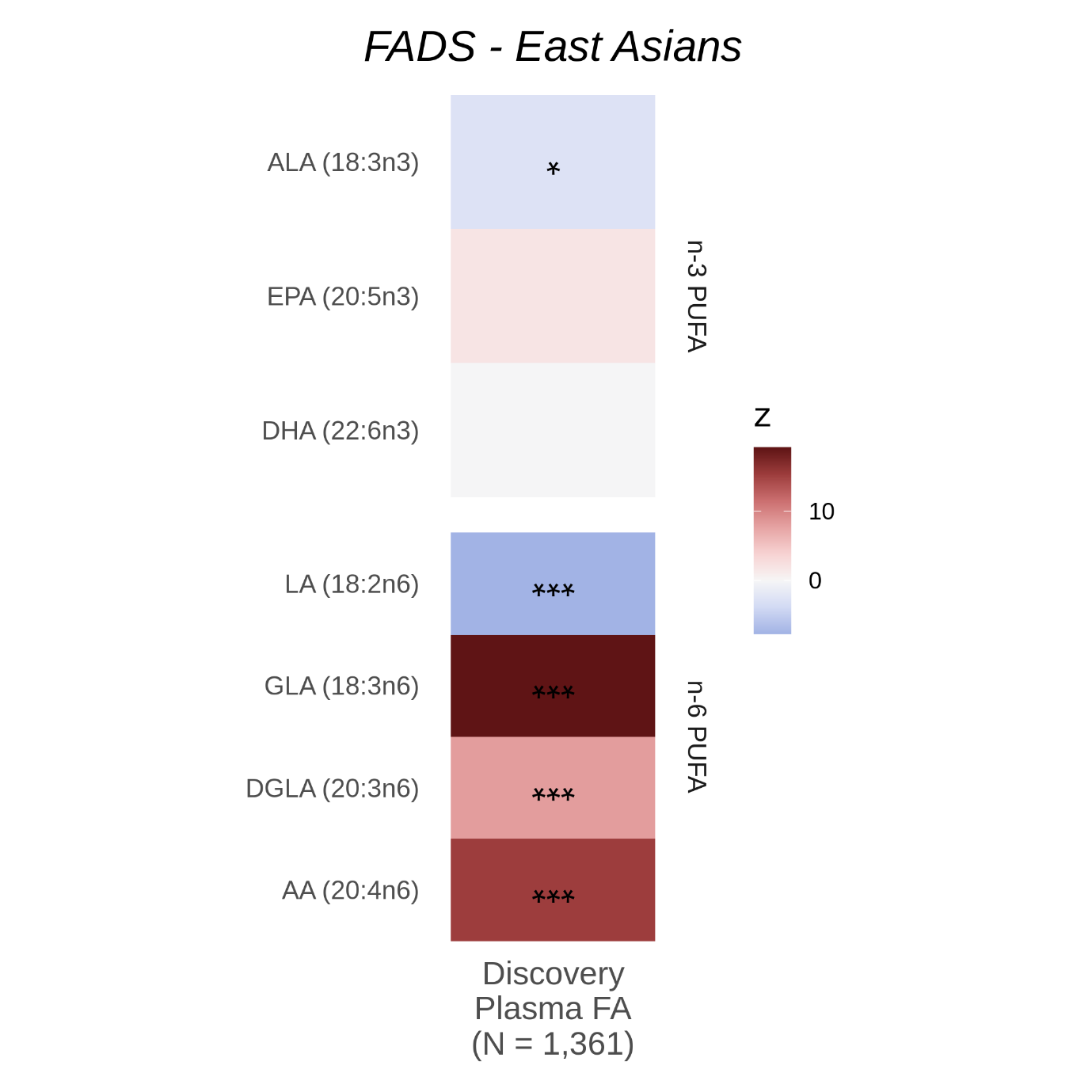

**Supplementary figure 3**

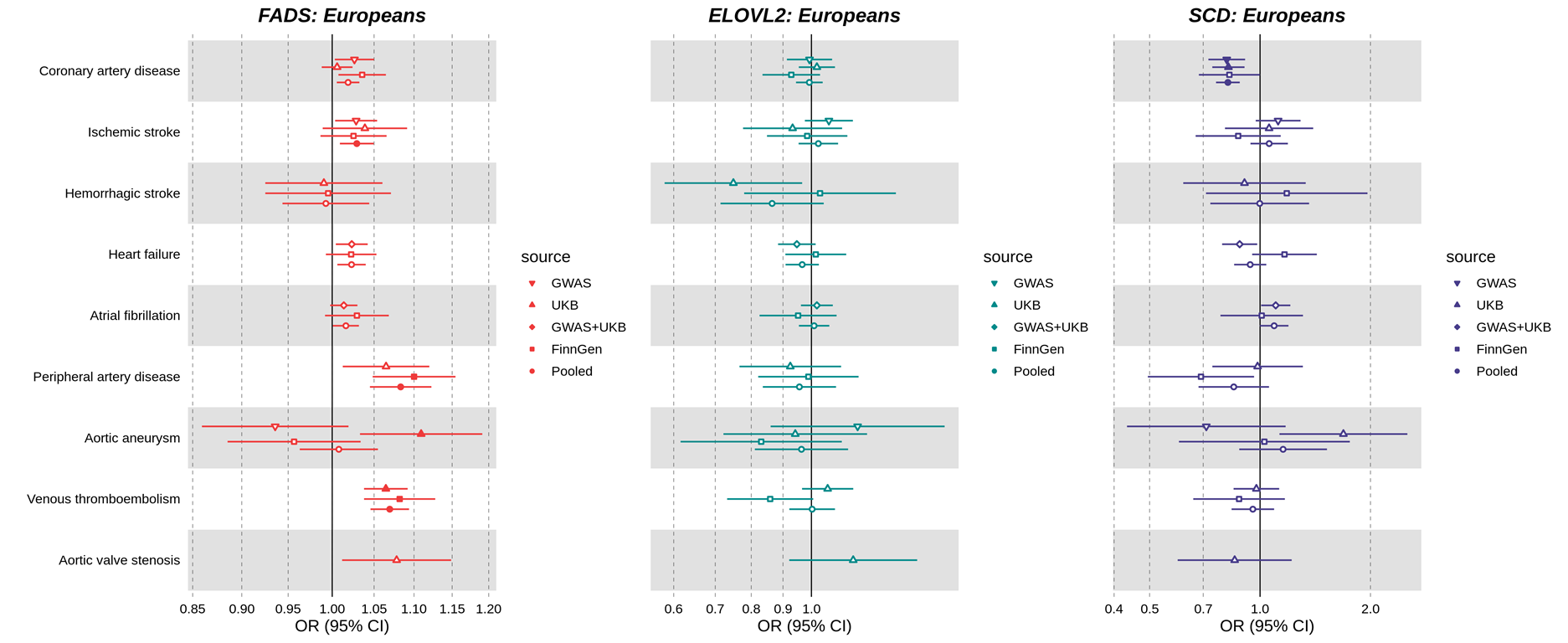

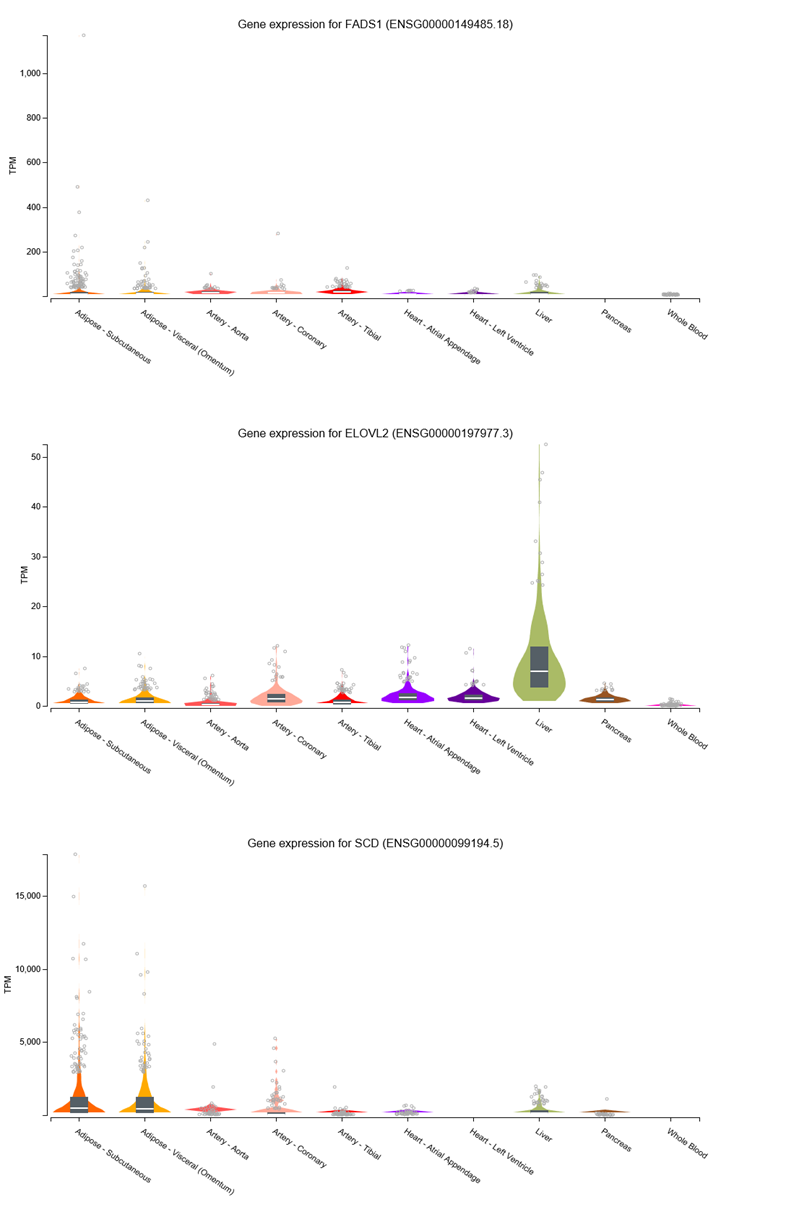

**Supplementary figure 4**

**Supplementary figure 5A**

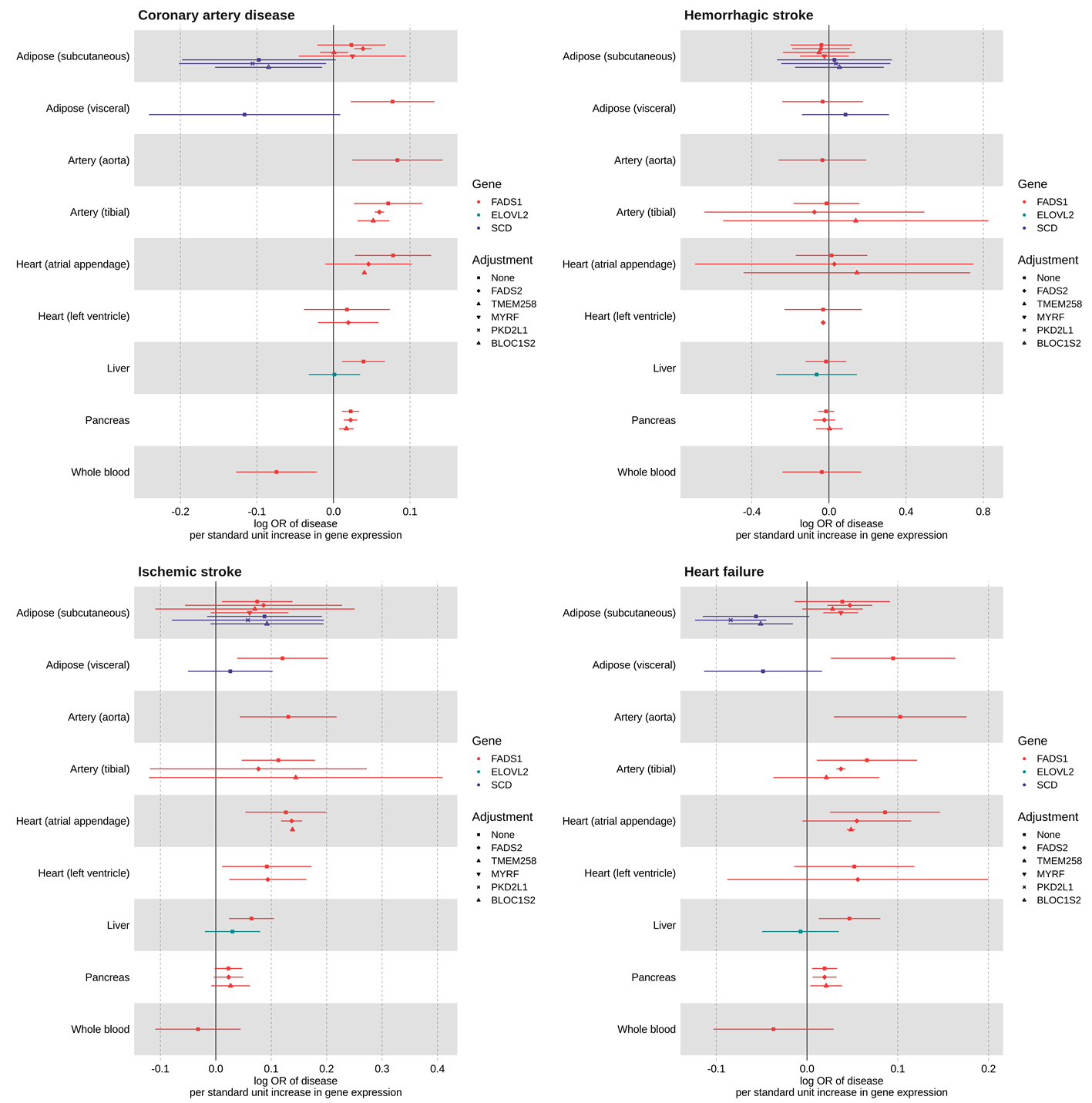

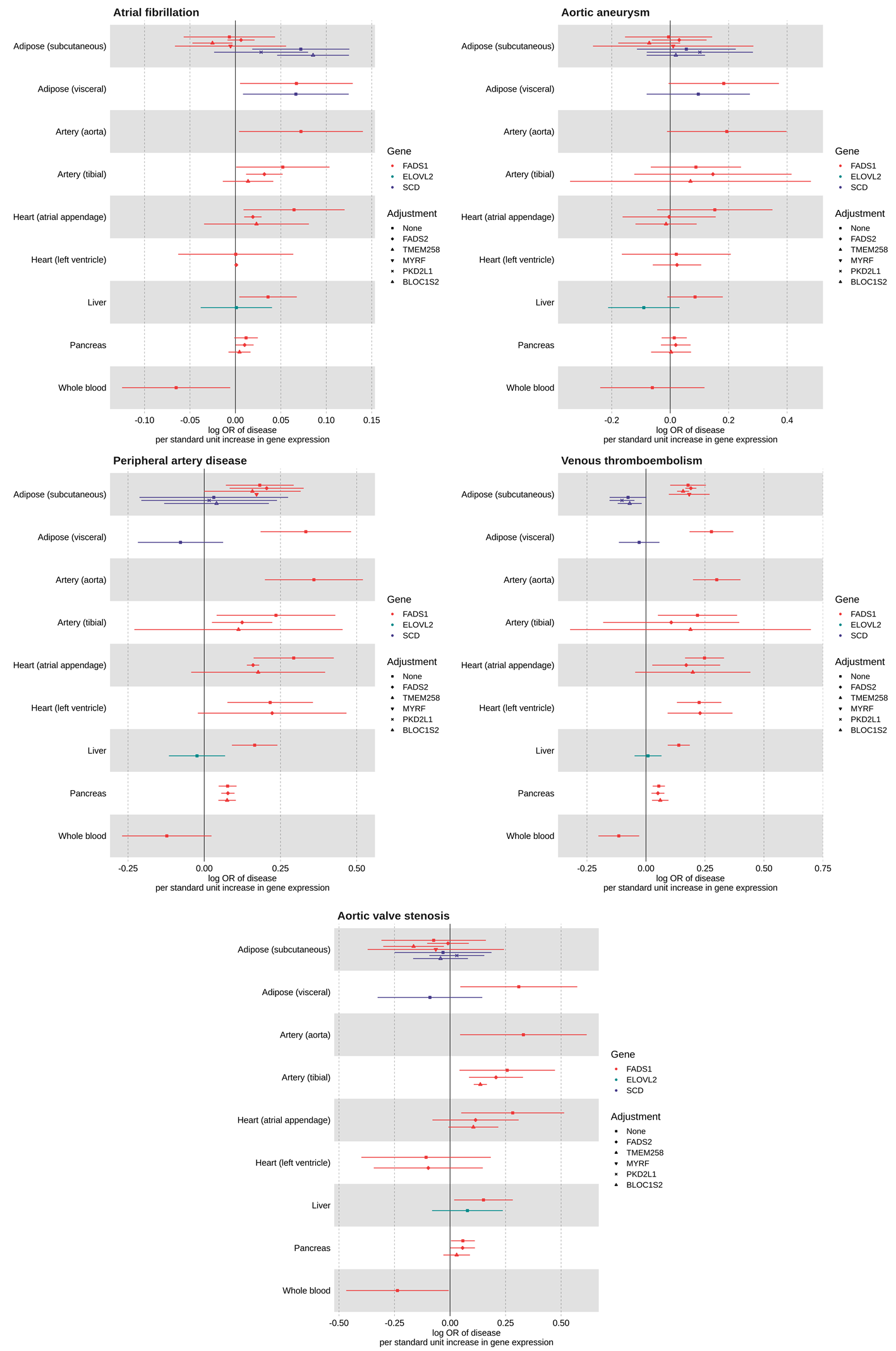

**Supplementary figure 5B**

**Supplementary figure 6A**

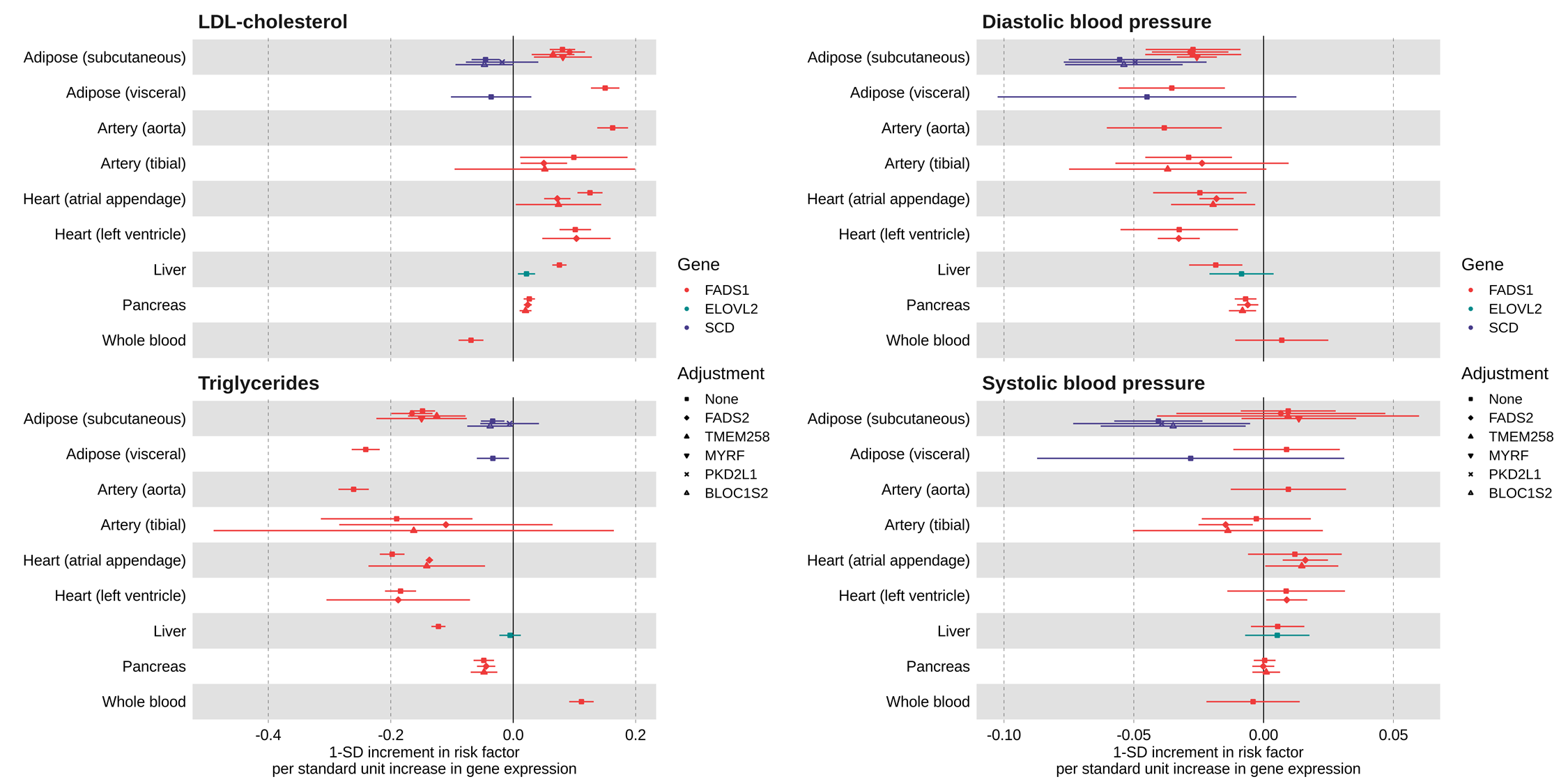

**Supplementary figure 6B**

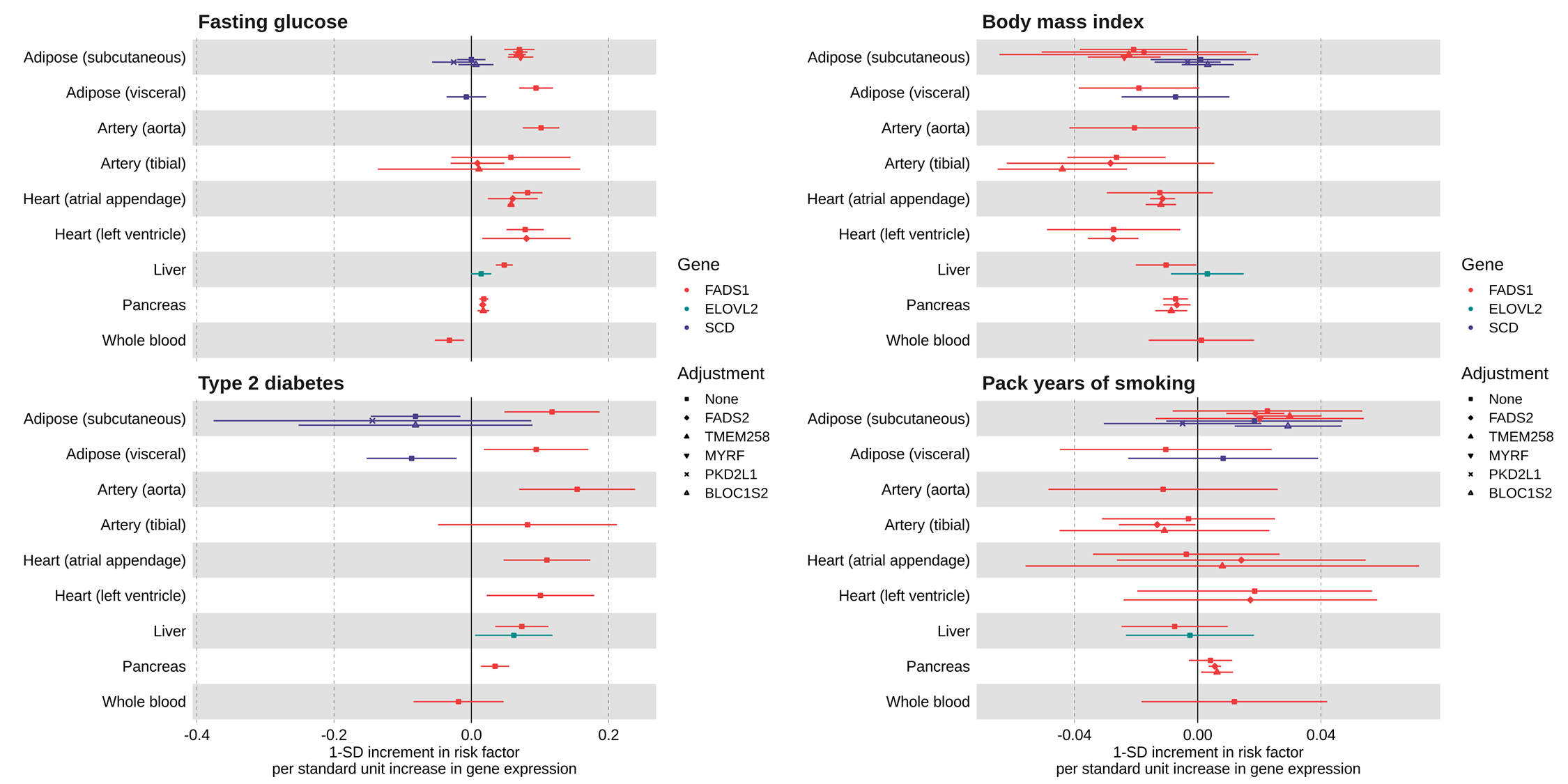

**Supplementary figure 7**

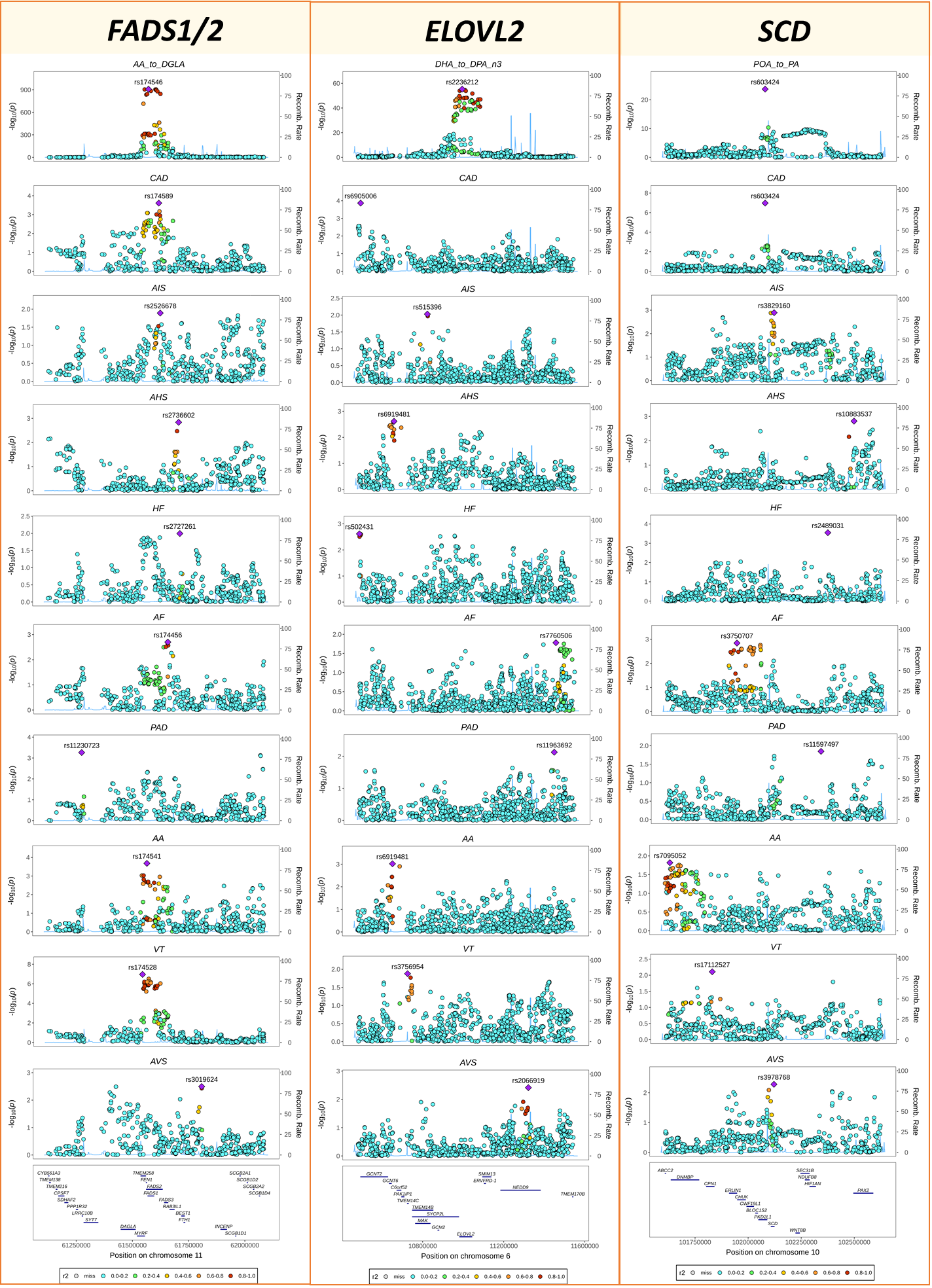

**Supplementary figure 8**

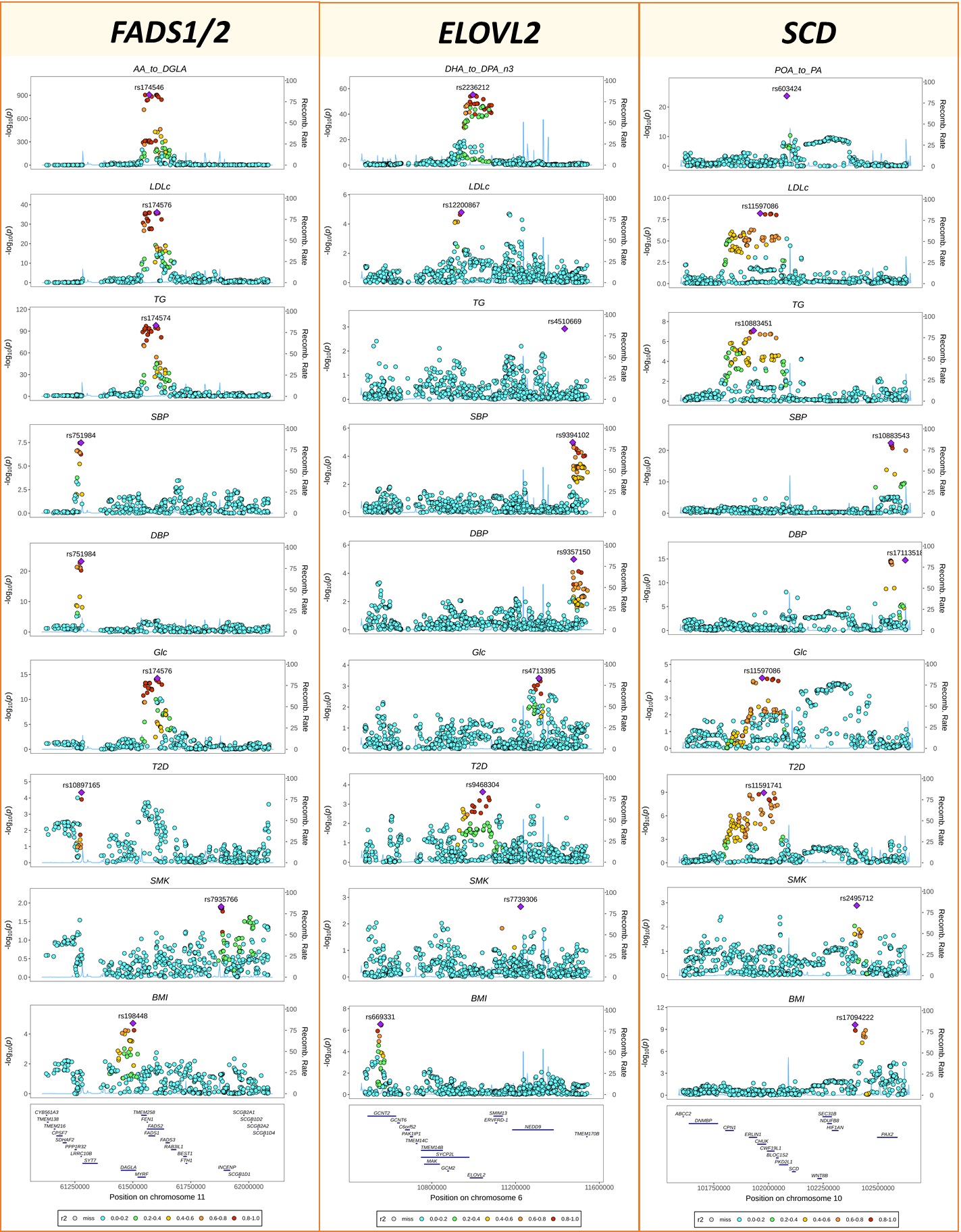

**Supplementary figure 9**

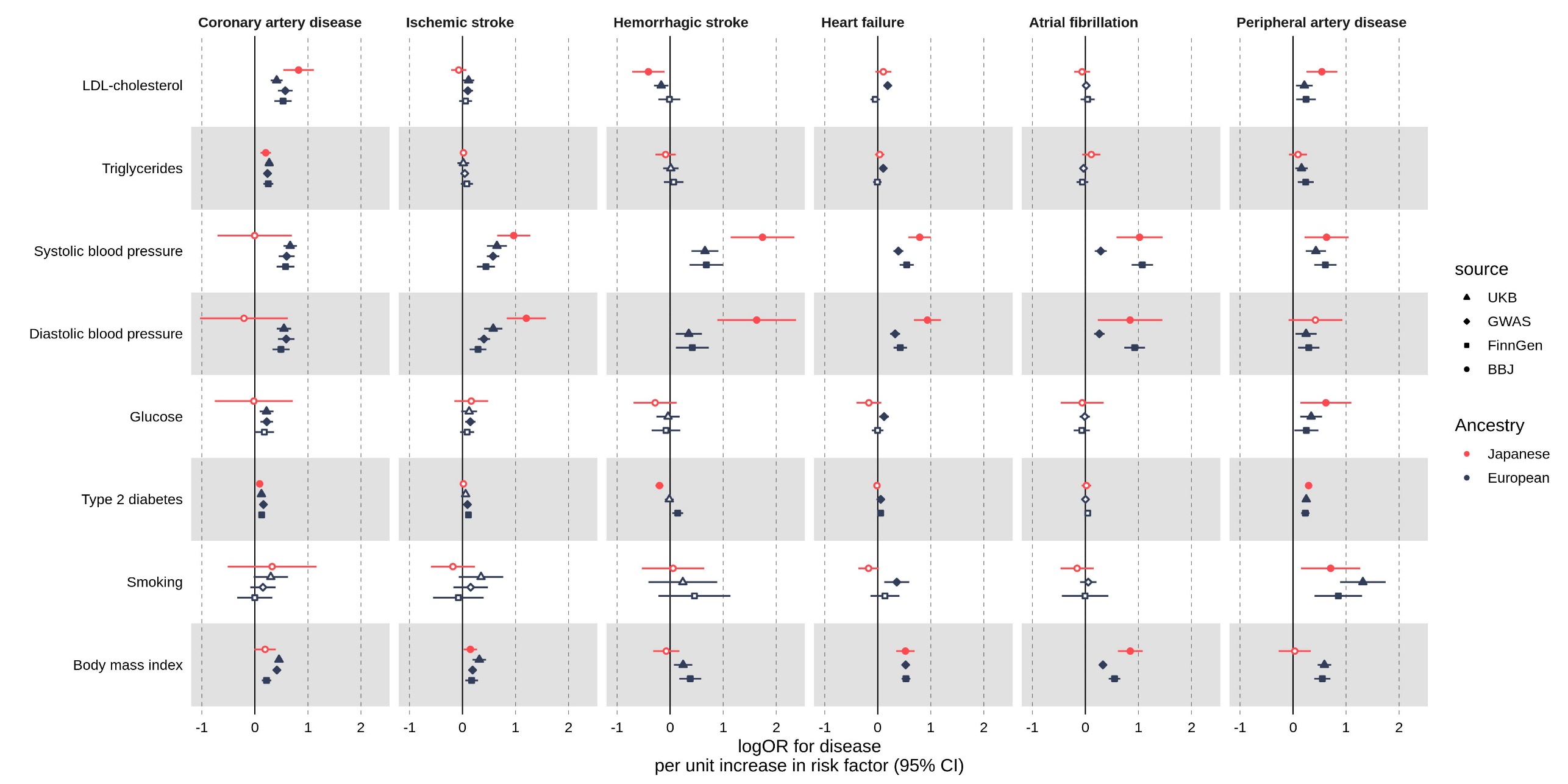
